# Joint representations from multi-view MRI-based learning support cognitive and functional performance domains

**DOI:** 10.1101/2025.09.27.25336706

**Authors:** Brian B Avants, Nicholas J Tustison, James R Stone

## Abstract

**Background:** Multimodal MRI (sMRI, dMRI, rsfMRI) encodes complementary aspects of brain structure and function; principled joint representations promise more sensitive and interpretable markers of brain health than single-modality features.

**Methods:** We evaluate Normative Neurological Health Embedding (NNHEmbed), a flexible multi-view framework that uses constrained cross-modal similarity objectives to learn low-dimensional embeddings. Models were trained and tested on the UK Biobank (n = 21,300) and evaluated for transfer and longitudinal sensitivity in independent cohorts (Normative Neuroimaging Library, Alzheimer’s Disease Neuroimaging Initiative, Parkinson’s Progression Markers Initiative).

**Results:** NNHEmbed produced compact, biologically interpretable components that (a) maps to established neurocognitive systems (e.g., episodic memory, processing-speed, sensorimotor/basal-ganglia circuits), (b) generalizes across cohorts, and (c) captures within-subject change over time. Best configurations balance reconstruction fidelity and shared covariance, improving interpretability while preserving predictive utility. Case demonstrations illustrate individualized normative profiling across multiple visits.

**Conclusions:** NNHEmbed yields stable, transferable multimodal embeddings suitable for normative mapping and longitudinal monitoring. Software, NNHEmbed configurations and derived bases are available for reproduction and reuse.

## 1 Introduction

The human brain is organized hierarchically, with its 86 billion neurons (Herculano-Houzel 2009) clustered into a few hundred cytoarchitectonically distinct regions (Amunts et al. 2020). These regions serve as computational units, defined by their characteristic cell types, laminar organization, signaling dynamics, and long-range connectivity profiles. Increasing biological and computational evidence suggests that interactions across these levels of organization give rise to low-dimensional neural representations (Gao and Ganguli 2015; Gallego et al. 2017). Such representations support efficient information processing, enabling flexible cognitive operations across functional domains including perception, language, and motor control (Shine et al. 2019).

Identifying the shared organizational principles of the human brain that underlie cognition remains a central challenge in cognitive neuroscience. Multi-modality magnetic resonance imaging (M3RI) provides a uniquely powerful framework for this endeavor, as it enables the concurrent characterization of cortical and subcortical anatomy (sMRI), white matter microstructure (dMRI), and large-scale functional connectivity (rsfMRI). Together, these modalities offer a complementary window onto the mechanisms of structure–function coupling in vivo (Calhoun and Sui 2016; Sui et al. 2020; Sporns 2005; Glasser et al. 2016). The integration of such diverse measurements is strongly motivated by foundational principles in systems neuroscience, which establish that the brain’s functional architecture is inextricably shaped and constrained by its underlying anatomical substrate (Ramón y Cajal 1899; Mountcastle 1997; Passingham, Stephan, and Kötter 2002). Crucially, multivariate and fusion-based methods that capture the joint patterns across modalities move beyond the constraints of single-measure analyses, yielding a more holistic and biologically grounded account of how structural and functional features converge to support cognition.

Multi-view learning methods are ideally suited for this task when it is approached in a data-driven manner. Such methods take advantage of complementary information between modalities to enhance sensitivity and robustness. Methods such as multi-set canonical correlation analysis (mCCA) and joint independent component analysis (jICA) (Meng et al. 2016; Anowar, Sadaoui, and Selim 2021) have revealed distinct structure-function disruptions in neurological and psychiatric disorders including schizophrenia, Parkinson’s disease, major depressive disorder, and Alzheimer’s disease (Sui et al. 2015; Groves et al. 2011). Beyond clinical populations, multimodal fusion has proven invaluable for characterizing normative brain variability across the lifespan, particularly when combined with behavioral, cognitive, and genetic data (Karlaftis et al. 2019; Marek, Tervo-Clemmens, et al. 2022; Elliott et al. 2018). As neuroimaging datasets become both deeper (larger sample sizes) and wider (more individual variables), integrative methods have become increasingly viable and even necessary for modeling the associated scientific questions.

Within the framework of multi-view methods, techniques that emphasize sparse, multivariate feature extraction are uniquely suited for uncovering interpretable latent patterns that only emerge when modalities are studied in concert (Sui et al. 2020; Mihalik et al. 2022). Our objective is to evaluate one such integrative framework: Normative Neurological Health Embedding (NNHEmbed) which is a neuroimaging-specific derivative of Similarity-driven Multiview Linear Reconstruction (SiMLR) (Brian B. Avants et al. 2021) customized for automated imaging data phenotype (IDP) processing and normative modeling. NNHEmbed provides: biological specificity through interpretable features; statistical power through dimensionality reduction; and the ability to probe and quantify structure-function prediction of cognitive or behavioral outcomes (Smith et al. 2015; Cole 2020). NNHEmbed is well-suited to tasks wherein biological and statistical dependencies coexist across modalities. In M3RI, this directly reflects the notion that structural properties influence function and functional states are enabled by structure. NNHEmbed also provides clear feature-level interpretability within a multimodal context allowing researchers to identify precisely which regions, cortical measures, white matter tracts, or functional networks within a given modality are most relevant to a given latent representation and in predicting a given outcome. This sparse and specific solution space is far more interpretable, both qualitatively and quantitatively, than a single, merged loading matrix derived from concatenated views of data (Shamy et al. 2011; McKeown et al. 1998; Calhoun et al. 2001).

Despite the clear motivation behind methods like NNHEmbed, there remains a lack of large-scale empirical evaluation of such frameworks in normative populations. Most prior applications have focused on clinical diagnostics or disorder classification, leaving a gap in understanding how multi-view methods can be used to study cognitive and functional variability across the general population (Cole 2020; Ivanova et al. 2021). The growing availability of large-scale resources such as the UK Biobank (UKB) (Miller et al. 2016) and the Normative Neuroimaging Library (NNL) (Gage et al. 2024) offers new opportunities to systematically examine how structural, functional, and white matter organization jointly relate to cognition and behavior across the adult lifespan.

The UK Biobank stands out as a uniquely comprehensive resource, offering tens of thousands of participants with sMRI, fMRI, and dMRI linked to detailed phenotypic and behavioral assessments. Multimodal analyses using UKB have shown that integrating across imaging modalities leads to reliable associations with cognition, aging, and disease risk (Smith et al. 2020; Cole 2020). However, a key challenge to demonstrating utility of multi-view learning for brain-behavior association is the availability and consistency of the data not only within but also across M3RI cohorts. The UKB, for instance, provides an enormous resource of data but relatively shallow cognitive phenotyping. The NNL, on the other hand, provides high quality cognitive phenotyping across a broad range of ages but lacks the enormous sample sizes of UKB. The Alzheimer’s Disease Neuroimaging Initative (ADNI) and Parkinson’s Progression Markers Initiative (PPMI) represent populations that are not only healthy controls but also at-risk for or diagnosed with neurodegenerative disease. These cohorts provide both cross-sectional and longitudinal M3RI accompanied by study-specific cognitive and functional measures.

In this work, we seek to address these limitations by stacking and linking evaluation opportunities across these datasets. We thus train NNHEmbed on M3RI data from a cross-sectional subset of UKB. We then validate the learned structure-function patterns in test (cross-sectional) and a separate longitudinal partition of the UKB in order to further characterize the impact of objective function choice. We then evaluate NNHEmbed-derived patterns in complementary participant studies in order to demonstrate that patterns learned from largescale cohorts are generalizable. Lastly, we demonstrate the best-performing NNHEmbed incarnation as a normative model of neurological health (normative neurological health, NNH) using a held-out longitudinal sub-population.

The contributions of this study include:

1. Data-driven evaluation of the NNHEmbed parameter space for deriving compact, interpretable and biologically relevant representations from multi-modal MRI;
2. Presentation of the reusability, characterization and hierarchical interpretations of the latent representations that are derived from applying NNHEmbed;
3. Demonstration of how pre-trained, reusable multi-view bases can be applied across datasets to capture stable brain-behavior associations in cognitive and functional domains.

By adopting a training and testing framework and leveraging data from multiple healthy and clinical populations, we contribute to the field of neuroimaging by clarifying the role of multi-view feature learning in scalable, reproducible brain-behavior modeling. Additionally, we demonstrate the sensitivity of these learned patterns to longitudinal change at the individual level and thus their potential for normative assessments. All tools are open-source via the ANTsR platform (Brian B. Avants et al. 2021). Table G provides a glossary of terms used in this document.

## 2 Results

Figure 1 illustrates the overall NNHEmbed learning framework, while Figure 2 summarizes the evaluation metrics employed to compare different NNHEmbed configurations. Below, we present an overview of the evaluation results.

**Figure 1.**
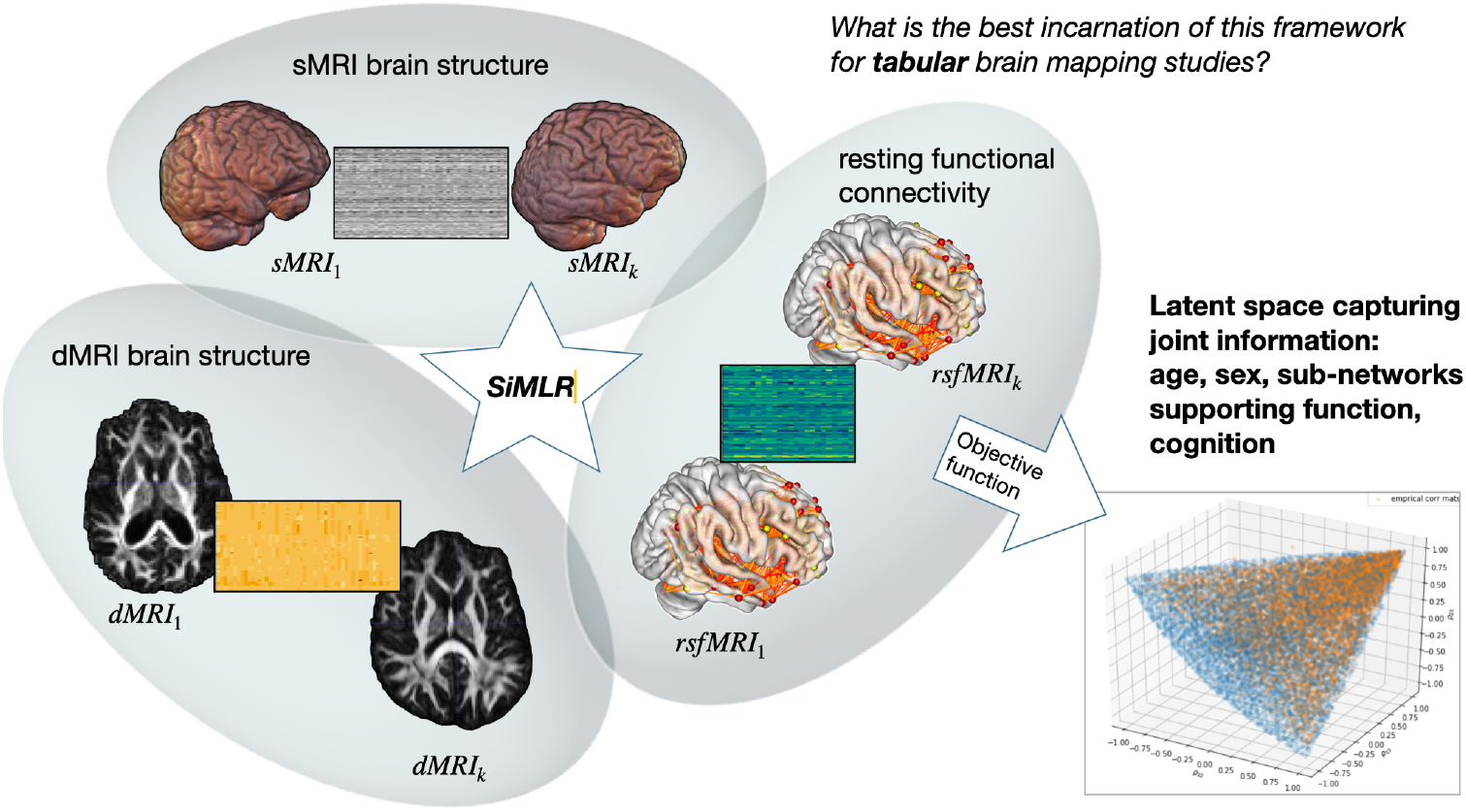
The overall learning framework used across NNHEmbed configurations. The approach integrates multiple MRI modalities (e.g., structural, diffusion, resting-state) into a shared low-dimensional representation by optimizing different objective functions (e.g., reconstruction fidelity, cross-modal covariance). Each configuration reflects a unique balance between capturing modality-specific variance and isolating shared variance across modalities. The resulting embeddings serve as a basis for brain–behavior mapping, cross-cohort transfer, and longitudinal prediction of cognitive and clinical outcomes.

**Figure 2.**
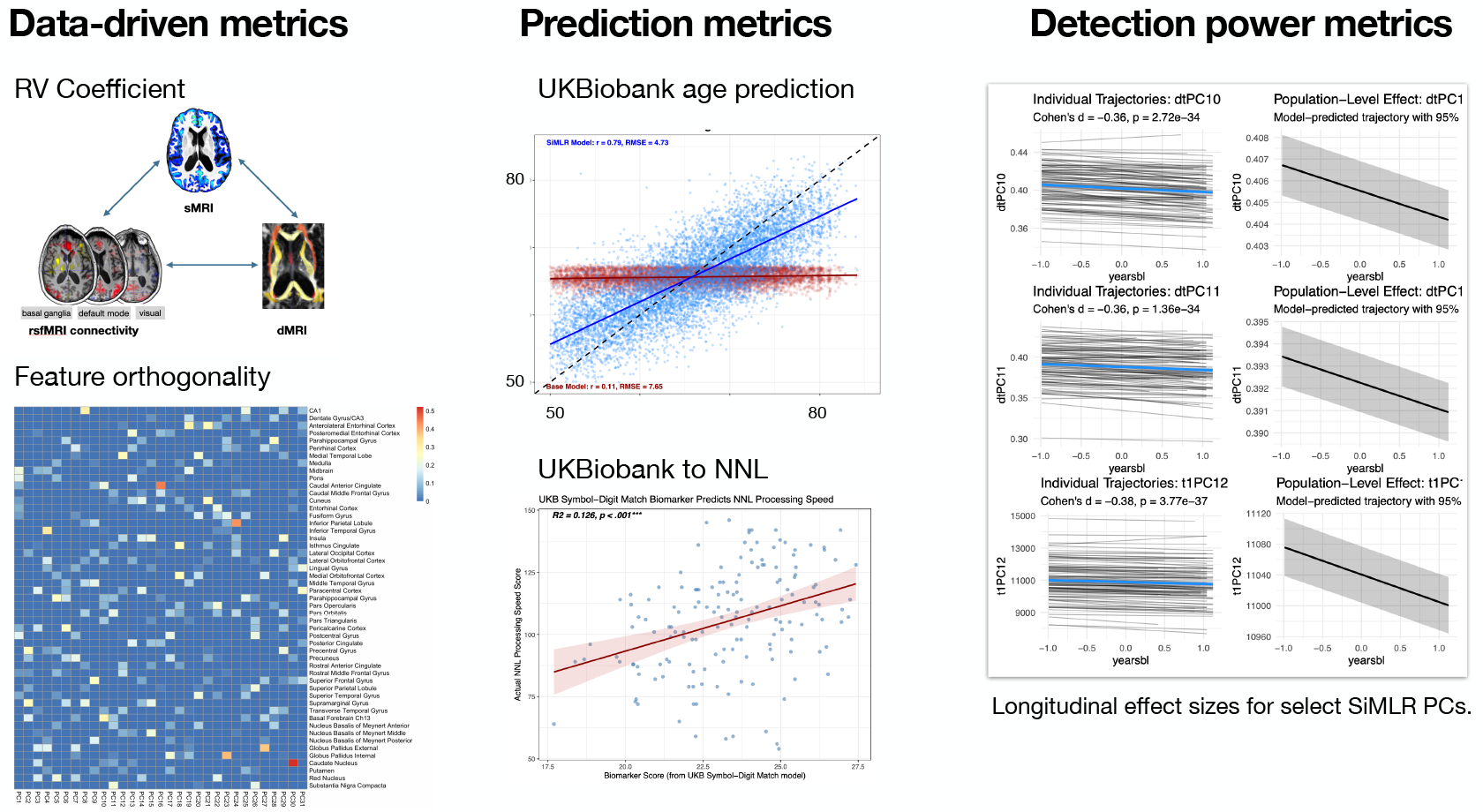
Evaluation metrics used to compare NNHEmbed configurations. Multiple complementary criteria were employed to assess the quality and utility of the learned embeddings. These included measures of representation quality, cross-modal alignment (e.g., shared covariance across modalities), and behavioral relevance (e.g., predictive accuracy for cognitive and clinical outcomes). Longitudinal sensitivity was also considered, evaluating whether the embeddings captured within-subject change over time. Together, these metrics provide a comprehensive basis for selecting the most effective NNHEmbed configuration for brain–behavior mapping.

### 2.1 Overview of evaluation results

We aggregate evaluation results from 144 NNHEmbed configurations run on tabular representations of M3RI and additional baseline methods for reference. Baseline methods include PCA, sparse generalized CCA (Tenenhaus and Tenenhaus 2011), regularized generalized CCA (Tenenhaus and Tenenhaus 2011) and an implementation of the original generalized CCA method (KETTENRING 1971). All methods used a basis size of 31 which was automatically derived from analysis of the scree plots of individual modality tabular matrices; 31 was the minimum basis size that captures 95% of the variance in an individual modality and, as such, acts as an upper bound on the size of the joint representation (see Appendix 1). Overall ranking derived from weighted objective data-driven metrics (weight 0.25) and application-driven metrics are shown in Table 1. Objective data-driven metric include feature orthogonality and the RV coefficient (Robert and Escoufier 1976) (see Table 2). Three separate predictions tasks are included: 1. age prediction in UKB train to UKB test (weight 0.25); 2. longitudinal sensitivity to change in brain measurements in the UKB longitudinal test set (weight 1.0); 3. cross-dataset cognitive prediction from UKB to NNL as shown in Table 3 (weight 1.0). We provide these latter tasks with the strongest weight as they are the most challenging and important in this context. That is, we want the methodology to both capture change in the brain over time and also provide a generalizable and predictive representation.

**Table 1:** Evaluation results. Top 20 of 145. The baseline PCA method is ranked 80th. Method names encode, in shorthand, the similarity measure (acc = absolute canonical covariance; osh = logcosh; reg = regression; nc = normalized correlation), mixing method (svd, PCA, ICA) and constraints used in the configuration (dom and corth are domain knowledge priors and orthogonality constraints respectively).

| Rank | Method | UKB.RV | F.Orth | UKB.Lo | UKB.NNL | f2.AD.PD.NNL |
| --- | --- | --- | --- | --- | --- | --- |
| 1 | acc.ica.dom.0.corth0.5x100.n18 | 0.072 | 0.007 | 0.376 | 0.132 | 31.127 |
| 2 | nc.pca.dom.1.corth0.1x100.n97 | 0.067 | 0.010 | 0.382 | 0.124 | 28.526 |
| 3 | nc.ica.dom.1.corth0.5x100.n88 | 0.072 | 0.006 | 0.379 | 0.119 | 29.043 |
| 4 | nc.ica.dom.0.corth0.9x100.n96 | 0.073 | 0.007 | 0.396 | 0.118 | 29.715 |
| 5 | nc.svd.dom.1.corth0.1x100.n73 | 0.075 | 0.007 | 0.379 | 0.118 | 29.372 |
| 6 | nc.pca.dom.0.5.corth0.1x100.n98 | 0.067 | 0.009 | 0.363 | 0.126 | 24.579 |
| 7 | nc.pca.dom.0.corth0.1x100.n99 | 0.072 | 0.009 | 0.366 | 0.119 | 29.571 |
| 8 | acc.ica.dom.1.cnone.n19 | 0.083 | 0.067 | 0.365 | 0.122 | 29.539 |
| 9 | acc.svd.dom.0.corth0.1x100.n3 | 0.076 | 0.011 | 0.351 | 0.128 | 29.477 |
| 10 | osh.pca.dom.0.corth0.1x100.n135 | 0.127 | 0.232 | 0.367 | 0.121 | 35.283 |
| 11 | nc.svd.dom.1.corth0.5x100.n76 | 0.076 | 0.004 | 0.348 | 0.126 | 29.670 |
| 12 | nc.pca.dom.0.5.corth0.5x100.n101 | 0.072 | 0.007 | 0.366 | 0.117 | 29.560 |
| 13 | acc.ica.dom.0.5.corth0.5x100.n17 | 0.083 | 0.012 | 0.346 | 0.129 | 31.481 |
| 14 | osh.svd.dom.0.5.corth0.9x100.n119 | 0.097 | 0.043 | 0.364 | 0.118 | 33.304 |
| 15 | nc.svd.dom.0.corth0.1x100.n75 | 0.073 | 0.009 | 0.346 | 0.128 | 27.079 |
| 16 | osh.ica.dom.1.corth0.1x100.n121 | 0.110 | 0.123 | 0.391 | 0.116 | 32.240 |
| 17 | osh.svd.dom.1.corth0.9x100.n118 | 0.100 | 0.067 | 0.363 | 0.118 | 35.242 |
| 18 | osh.pca.dom.0.5.corth0.5x100.n137 | 0.128 | 0.238 | 0.369 | 0.117 | 34.437 |
| 19 | reg.ica.dom.0.corth0.5x100.n54 | 0.066 | 0.008 | 0.350 | 0.126 | 29.721 |
| 20 | osh.pca.dom.0.corth0.5x100.n138 | 0.125 | 0.229 | 0.371 | 0.117 | 37.035 |
| 35 | base.pca.cnone.n145 | 0.064 | 0.000 | 0.210 | 0.127 | 17.760 |

**Table 2:**
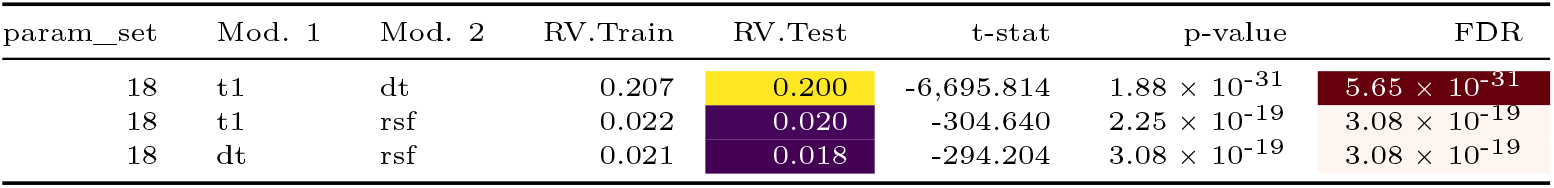
Pairwise RV Coefficient Analysis with *t*-test (lower better) and *p*-value assessed by permutation.

**Table 3:** Data from UKB Symbol-Digit Match to Predict NNL Proc.Speed.

| Term | Estimate | Std. Error | Statistic | p-value | 95% CI |
| --- | --- | --- | --- | --- | --- |
| (Intercept) | 25.841 | 18.711 | 1.381 | 0.169 | [-11.12, 62.8] |
| predicted_biomarker | 3.648 | 0.852 | 4.281 | 0.000 | [1.96, 5.33] |

*Model Fit Statistics:* R-squared = 0.105 ; Adj. R-squared = 0.099 ; n = 159

The evaluation thus balances the diversity of the features themselves (feature orthogonality), performance on multi-view inter-modality association (the RV coefficients measured between the M3RI embeddings), cross-cohort predictive accuracy (UK Biobank processing speed to a related processing speed metric in the NNL (Carlozzi et al. 2015)) and longitudinal detection power indicators (Cohen’s *d* for the multivariate mixed effects model improvement provided by the interaction of embedding variables with time measured as years from baseline). By applying the `rank_methods_by_performance` function from the `R` package `subtyper` (Brian B. Avants 2025), we obtained an overall ordering of the NNHEmbed configurations and the baseline methods. The resulting ranks, shown in Table 1, highlight which configurations achieve superior performance. Additional details of these evaluation strategies and the results are discussed in the methods section. Table 4 shows the demographic characteristics of the UK Biobank training set. Table 5 details the UKB test set. Table 6 shows the UKB longitudinal set (completely unique subjects from the other divisions). Table 7 shows the NNL test set demographics.

**Table 4.** UKB training set population characteristics.

| Characteristic | Female N = 4,491<sup>†</sup> | Male N = 3,870<sup>†</sup> |
| --- | --- | --- |
| **age.mri** | 65 (59, 71) | 67 (60, 72) |
| **ethnic.background.f2** |  |  |
| African | 9 (0.2%) | 9 (0.2%) |
| Any other Asian background | 2 (<0.1%) | 10 (0.3%) |
| Any other mixed background | 3 (<0.1%) | 5 (0.1%) |
| Any other white background | 196 (4.4%) | 99 (2.6%) |
| Bangladeshi | 0 (0%) | 1 (<0.1%) |
| British | 4,041 (90%) | 3,543 (92%) |
| Caribbean | 22 (0.5%) | 12 (0.3%) |
| Chinese | 17 (0.4%) | 2 (<0.1%) |
| Do not know | 2 (<0.1%) | 0 (0%) |
| Indian | 19 (0.4%) | 33 (0.9%) |
| Irish | 123 (2.7%) | 107 (2.8%) |
| Other ethnic group | 29 (0.6%) | 19 (0.5%) |
| Pakistani | 2 (<0.1%) | 12 (0.3%) |
| Prefer not to answer | 5 (0.1%) | 14 (0.4%) |
| White | 3 (<0.1%) | 1 (<0.1%) |
| White and Asian | 8 (0.2%) | 2 (<0.1%) |
| White and Black African | 5 (0.1%) | 0 (0%) |
| White and Black Caribbean | 3 (<0.1%) | 1 (<0.1%) |
| Unknown | 2 | 0 |
| **educational.attainment** |  |  |
| A levels/AS levels or equivalent | 602 (14%) | 453 (12%) |
| College or University degree | 1,998 (45%) | 1,797 (47%) |
| CSEs or equivalent | 184 (4.2%) | 176 (4.6%) |
| None of the above | 290 (6.6%) | 259 (6.8%) |
| NVQ or HND or HNC or equivalent | 160 (3.6%) | 305 (8.0%) |
| O levels/GCSEs or equivalent | 917 (21%) | 647 (17%) |
| Other professional qualifications eg: nursing, teaching | 259 (5.9%) | 168 (4.4%) |
| Prefer not to answer | 16 (0.4%) | 18 (0.5%) |
| Unknown | 65 | 47 |
| **townsend.deprivation** | -2.54 (-3.83, -0.38) | -2.66 (-3.88, -0.60) |
| Unknown | 3 | 2 |
| **fluid.intelligence.sc** | 7.00 (5.00, 8.00) | 7.00 (5.00, 8.00) |
| Unknown | 3,040 | 2,596 |
| **DTI_dti_FD_mean** | 2.51 (2.15, 2.90) | 2.53 (2.16, 3.00) |
| **rsfMRI_fcnxpro122_FD_mean** | 0.14 (0.11, 0.18) | 0.16 (0.12, 0.21) |
| **T1Hier_resnetGrade** | 1.73 (1.48, 2.01) | 1.68 (1.43, 1.95) |
| **snr** | 10.09 (8.87, 11.35) | 10.45 (9.07, 11.82) |

<sup>†</sup>Median (Q1, Q3); n (%)

**Table 5.** UKB testing set population characteristics.

| Characteristic | Female N = 4,353<sup>i</sup> | Male N = 4,008<sup>i</sup> |
| --- | --- | --- |
| **age.mri** | 65 (59, 71) | 67 (61, 72) |
| **ethnic.background.f2** |  |  |
| African | 9 (0.2%) | 14 (0.3%) |
| Any other Asian background | 3 (<0.1%) | 9 (0.2%) |
| Any other Black background | 0 (0%) | 2 (<0.1%) |
| Any other mixed background | 12 (0.3%) | 6 (0.1%) |
| Any other white background | 175 (4.0%) | 93 (2.3%) |
| Bangladeshi | 0 (0%) | 1 (<0.1%) |
| British | 3,916 (90%) | 3,657 (91%) |
| Caribbean | 17 (0.4%) | 11 (0.3%) |
| Chinese | 16 (0.4%) | 8 (0.2%) |
| Indian | 18 (0.4%) | 40 (1.0%) |
| Irish | 123 (2.8%) | 108 (2.7%) |
| Mixed | 1 (<0.1%) | 0 (0%) |
| Other ethnic group | 28 (0.6%) | 23 (0.6%) |
| Pakistani | 4 (<0.1%) | 8 (0.2%) |
| Prefer not to answer | 10 (0.2%) | 15 (0.4%) |
| White | 1 (<0.1%) | 3 (<0.1%) |
| White and Asian | 9 (0.2%) | 3 (<0.1%) |
| White and Black African | 2 (<0.1%) | 2 (<0.1%) |
| White and Black Caribbean | 7 (0.2%) | 5 (0.1%) |
| Unknown | 2 | 0 |
| **educational.attainment** |  |  |
| A levels/AS levels or equivalent | 576 (13%) | 430 (11%) |
| College or University degree | 1,952 (46%) | 1,936 (49%) |
| CSEs or equivalent | 179 (4.2%) | 147 (3.7%) |
| None of the above | 299 (7.0%) | 276 (7.0%) |
| NVQ or HND or HNC or equivalent | 123 (2.9%) | 318 (8.0%) |
| O levels/GCSEs or equivalent | 921 (21%) | 702 (18%) |
| Other professional qualifications eg: nursing, teaching | 229 (5.3%) | 139 (3.5%) |
| Prefer not to answer | 5 (0.1%) | 13 (0.3%) |
| Unknown | 69 | 47 |
| **townsend.deprivation** | -2.61 (-3.88, -0.44) | -2.58 (-3.93, -0.39) |
| Unknown | 4 | 3 |
| **fluid.intelligence.sc** | 7.00 (5.00, 8.00) | 7.00 (5.00, 8.00) |
| Unknown | 2,942 | 2,664 |
| **DTI_dti_FD_mean** | 2.49 (2.16, 2.87) | 2.55 (2.19, 3.04) |
| **rsfMRI_fcnxpro122_FD_mean** | 0.14 (0.11, 0.19) | 0.16 (0.12, 0.21) |
| **T1Hier_resnetGrade** | 1.74 (1.49, 2.00) | 1.68 (1.43, 1.96) |
| **snr** | 10.17 (8.97, 11.42) | 10.35 (8.99, 11.72) |

<sup>i</sup>Median (Q1, Q3); n (%)

**Table 6.** UKB Longitudinal Population Characteristics.

| Characteristic | Female N = 2,398<sup>†</sup> | Male N = 2,180<sup>†</sup> |
| --- | --- | --- |
| **age.mri** | 62 (56, 68) | 64 (57, 70) |
| **ethnic.background.f2** |  |  |
| African | 6 (0.3%) | 7 (0.3%) |
| Any other Asian background | 0 (0%) | 1 (<0.1%) |
| Any other mixed background | 3 (0.1%) | 2 (<0.1%) |
| Any other white background | 65 (2.7%) | 57 (2.6%) |
| Bangladeshi | 2 (<0.1%) | 2 (<0.1%) |
| British | 2,218 (92%) | 2,007 (92%) |
| Caribbean | 11 (0.5%) | 3 (0.1%) |
| Chinese | 10 (0.4%) | 2 (<0.1%) |
| Do not know | 1 (<0.1%) | 0 (0%) |
| Indian | 7 (0.3%) | 24 (1.1%) |
| Irish | 45 (1.9%) | 53 (2.4%) |
| Other ethnic group | 15 (0.6%) | 6 (0.3%) |
| Pakistani | 4 (0.2%) | 5 (0.2%) |
| Prefer not to answer | 2 (<0.1%) | 6 (0.3%) |
| White | 1 (<0.1%) | 0 (0%) |
| White and Asian | 2 (<0.1%) | 3 (0.1%) |
| White and Black African | 0 (0%) | 1 (<0.1%) |
| White and Black Caribbean | 6 (0.3%) | 0 (0%) |
| Unknown | 0 | 1 |
| **educational.attainment** |  |  |
| A levels/AS levels or equivalent | 330 (14%) | 295 (14%) |
| College or University degree | 1,088 (46%) | 1,002 (47%) |
| CSEs or equivalent | 111 (4.7%) | 94 (4.4%) |
| None of the above | 109 (4.6%) | 103 (4.8%) |
| NVQ or HND or HNC or equivalent | 67 (2.8%) | 182 (8.5%) |
| O levels/GCSEs or equivalent | 522 (22%) | 386 (18%) |
| Other professional qualifications eg: nursing, teaching | 130 (5.5%) | 83 (3.9%) |
| Prefer not to answer | 5 (0.2%) | 3 (0.1%) |
| Unknown | 36 | 32 |
| **townsend.deprivation** | -2.57 (-3.87, -0.42) | -2.72 (-3.95, -0.68) |
| Unknown | 6 | 4 |
| **fluid.intelligence.sc** | 7.00 (5.00, 8.00) | 7.00 (5.00, 8.00) |
| Unknown | 1,602 | 1,416 |
| **DTI_dti_FD_mean** | 2.29 (1.99, 2.63) | 2.31 (2.01, 2.69) |
| **rsfMRI_fcnxpro122_FD_mean** | 0.13 (0.10, 0.17) | 0.15 (0.12, 0.20) |
| **T1Hier_resnetGrade** | 1.77 (1.51, 2.06) | 1.69 (1.45, 1.97) |
| **snr** | 10.03 (8.95, 11.26) | 10.55 (9.21, 11.98) |

<sup>†</sup>Median (Q1, Q3); n (%)

**Table 7:**
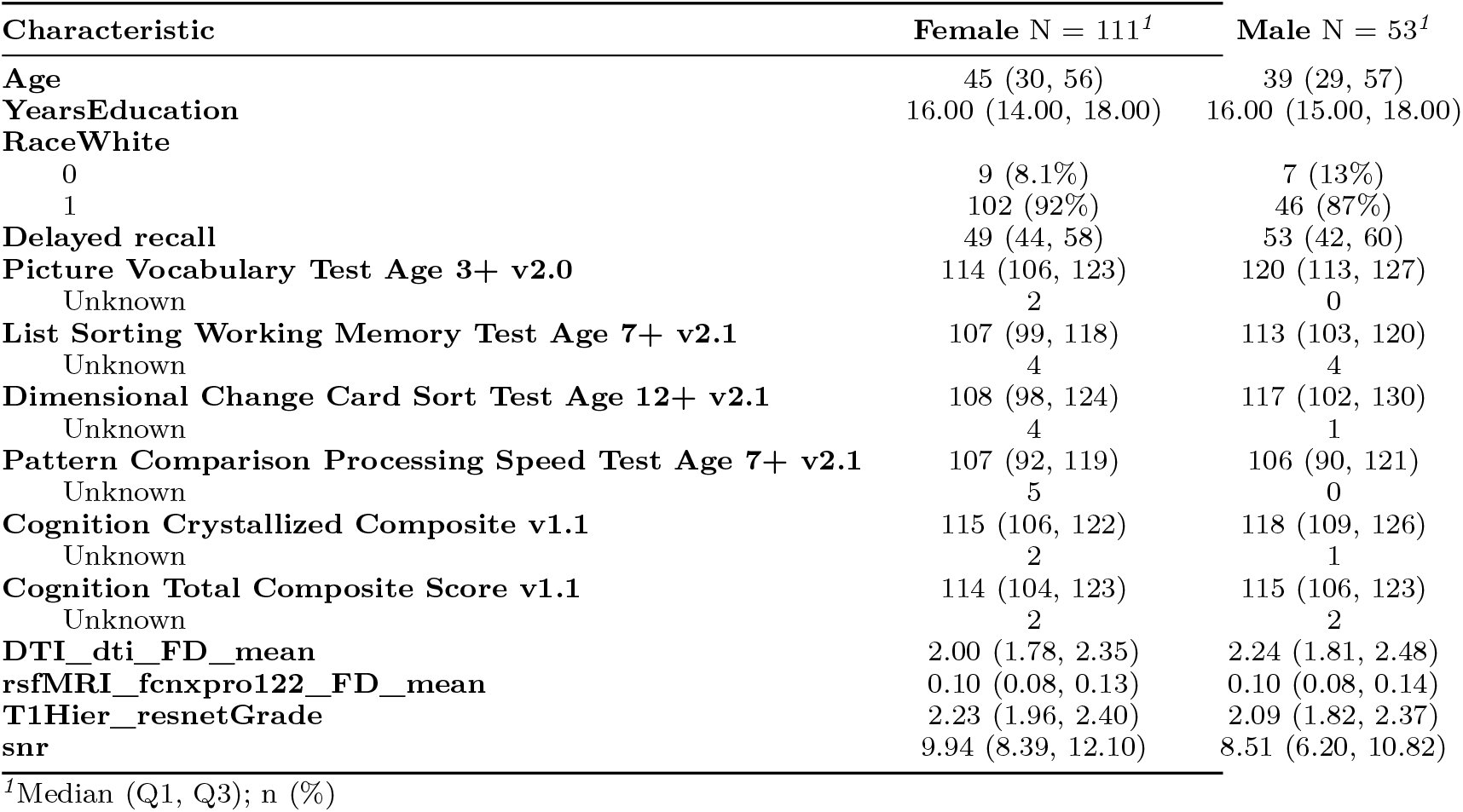
NNL Population Characteristics.

### 2.2 Results for best performing configuration

The best performing configuration, as determined by the weighted ranking of evaluation metrics, was acc.ica.dom.0.corth0.5x100.n18. See Table G for details of this shorthand (`n18` refers to the index of this parameter set). This configuration utilized absolute canonical covariance as the similarity measure, independent component analysis for identifying the mixed latent space in NNHEmbed, and incorporated moderate orthogonality constraints. The following sections present detailed results for this configuration, including inter-modality association analysis, cognitive prediction performance and longitudinal sensitivity in UKB. We also provide detailed neuroscientific interpretation of the learned features and apply these for tracking longitudinal brain changes in individual subjects.

#### 2.2.1 RV coefficient

The RV coefficient is a multivariate generalization of the Pearson correlation coefficient, measuring the similarity between two sets of variables. It ranges from 0 (no association) to 1 (perfect association). In this analysis, we compute the RV coefficients between pairs of imaging modalities in the UKB and compare the RV in the training data (on which NNHEmbed was computed) and the test data. This provides evidence that the results are not overfit to the training data. The results are further supported by comparing the coefficient values against that in permuted data and summarized in Table 2.

#### 2.2.2 Age prediction in the UKB test cohort

NNHEmbed embeddings were used to predict age in the UK Biobank test cohort. The results are summarized in Figure 3. These results use the full set of bases concatenated across modalities in a linear regression model (lm in R) trained in the UK Biobank training cohort. The results show good prediction accuracy, with a mean absolute error (MAE) of 3.922 years and an R-squared value of 0.593. The scatter plot illustrates the relationship between predicted and actual ages, indicating that the model captures age-related brain changes effectively. A typical MAE for well-regarded models is in the 3–5 year range. This includes models using either deep learning or more traditional machine learning approaches, often trained on publicly available datasets like UKB (Rokicki et al. 2021; Lange et al. 2022).

**Figure 3.**
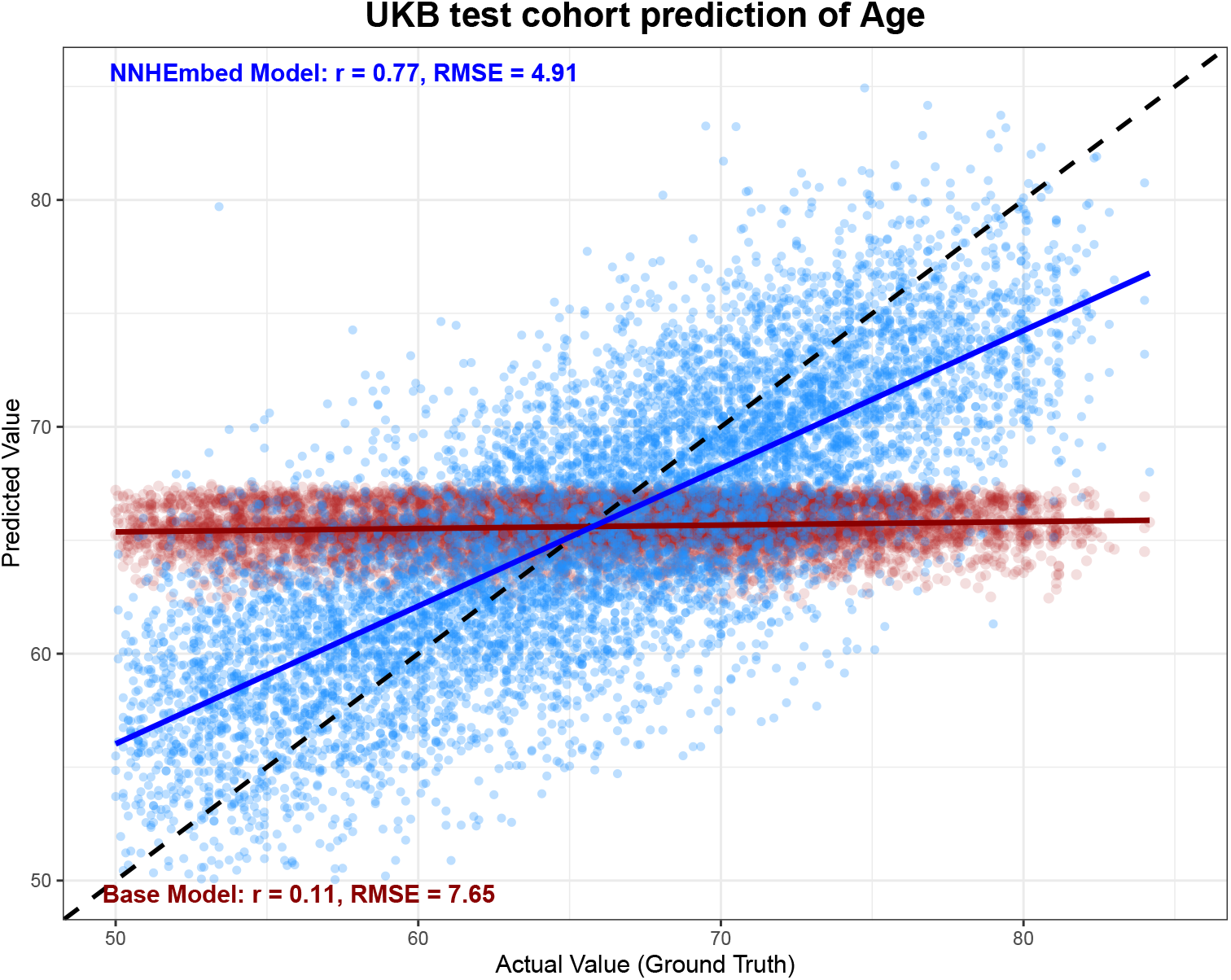
Age prediction in the UKB cohort from the best performing (overall) NNHEmbed embeddings. The base model uses only biological sex and the townsend deprivation index (TDI). The NNHEmbed model adds the full set of NNHEmbed embeddings from all modalities.

#### 2.2.3 Cross-cohort cognitive prediction

##### Cognitive Phenotype Training in UK Biobank (UKB)

A predictive NNHEmbed embedding-based ensemble model was developed using linear regression (`lm`). The models were trained on two separate data splits (UKB train and UKB test). The outcome variable for training was the “Symbol-Digit Match” test, a measure of processing speed. This measure was chosen because of the presence of closely related cognitive construct measurements in other data (a task from the National Institutes of Health (NIH) Toolbox-Cognitive Battery (NIHTB-CB) adjusted to account for a participant’s age (Carlozzi et al. 2015) and the ADNI Trails B measurement (Gibbons et al. 2012)). The trained linear models were used to generate a predicted pseudo-processing speed score for participants in two independent validation cohorts: ADNI and the NNL study. For each participant in these external cohorts, the final biomarker score was calculated as the average of the predictions from the two UKB-trained models. Only the NNL prediction score was used in ranking the NNHEmbed configurations (see Table 1); however, the ADNI prediction is included here for completeness. The results of these analyses in Figure 4 show that the UKB-derived biomarker is significantly associated with the harmonized cognitive outcomes in both validation cohorts, demonstrating its potential utility for cross-cohort cognitive prediction. Table 3 details regression results for the NNL prediction.

**Figure 4.**
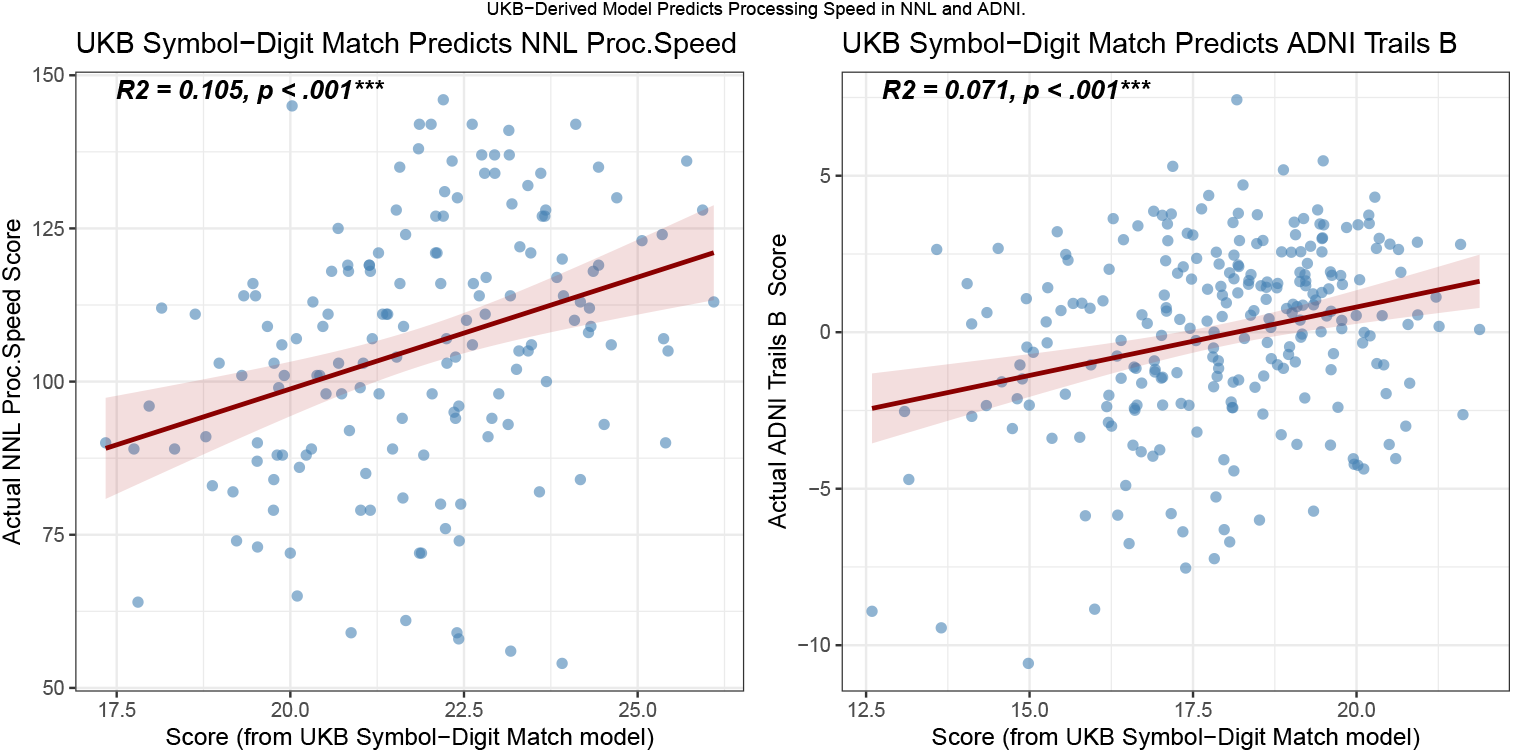
Cross-cohort cognitive prediction using the NNHEmbed-derived biomarker trained in UKB. The right panel shows the association in the ADNI cohort (mPACC Trails B score) and the left panel shows the association in the NNL cohort (NIH Toolbox Processing Speed score). The pseudo-biomarker is significantly associated with both outcomes, demonstrating its potential utility for cross-cohort cognitive prediction.

#### 2.2.4 Longitudinal detection power in the UKB longitudinal cohort

Longitudinal analysis of NNHEmbed embeddings in the UK Biobank longitudinal sample revealed modality-specific differences in sensitivity to brain change over time. All mixed-effects models included age, sex, and baseline brain volume as fixed effects, with participant ID entered as a random intercept. Effect sizes were summarized using Cohen’s *d* for the longitudinal (within-participant) component.

Across the 93 embeddings (31 per modality), **diffusion embeddings showed the strongest longitudinal effects**. Several dtPCs exceeded or approached moderate effect sizes, including dtPC2 (*d* = –0.622), dtPC5 (*d* = –0.547), dtPC13 (*d* = –0.528), dtPC28 (*d* = –0.563), dtPC25 (*d* = –0.513), dtPC6 (*d* = –0.414), dtPC11 (*d* = –0.476), dtPC9 (*d* = –0.366), and dtPC24 (*d* = –0.370). These findings indicate that the diffusion-derived latent features consistently captured aging-related microstructural decline.

Structural embeddings showed **moderate but generally smaller effects**, with the largest observed for t1PC31 (*d* = –0.397), t1PC7 (*d* = –0.402), t1PC2 (*d* = –0.364), t1PC6 (*d* = –0.364), t1PC12 (*d* = –0.309), and t1PC28 (*d* = –0.344). Although smaller than those from diffusion MRI, these structural embeddings still reflected measurable longitudinal variation in cortical and subcortical morphology.

In contrast, **resting-state fMRI embeddings showed uniformly weak longitudinal effects**. All rsfPCs were well below the |0.35| threshold, with the largest effect size observed for rsfPC10 (*d* = –0.218). This pattern suggests that the resting-state embeddings, as derived here, were less sensitive to aging-related within-person change over the available follow-up intervals.

For these significant embeddings, spaghetti and population-level plots (e.g., Figure 5) illustrated individual trajectories and model-predicted trends, confirming consistent declines with narrow confidence intervals. These findings highlight NNHEmbed’s ability to capture longitudinal changes in diffusion and structural modalities, supporting its utility in normative modeling for neurological health monitoring, particularly for detecting age-related declines.

**Figure 5.**
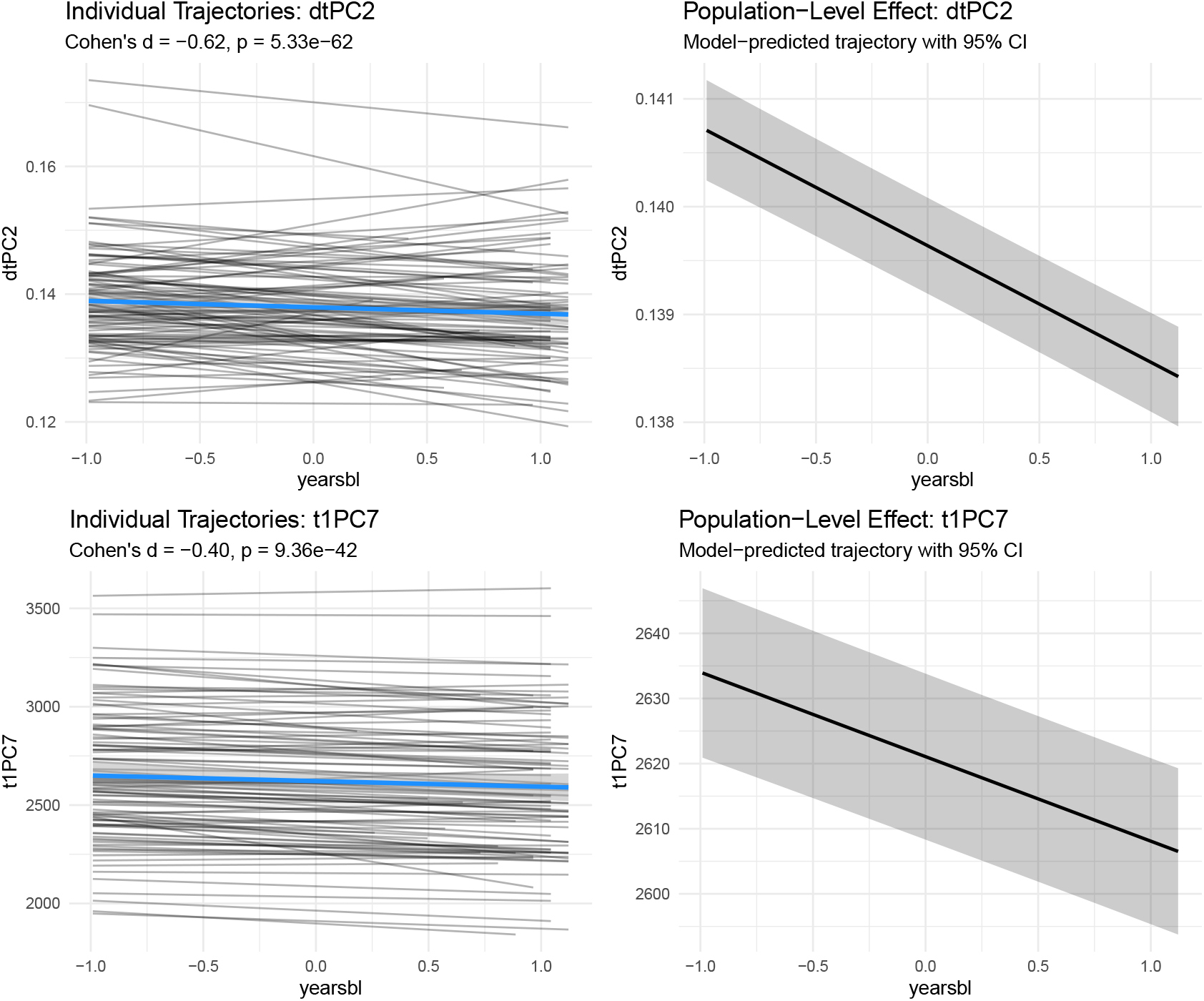
Longitudinal trajectories of NNHEmbed embeddings in the UK Biobank cohort for the most sensitive feature sets. Left panels show spaghetti plots of individual trajectories, and right panels display population-level trends with 95% confidence intervals, highlighting significant declines in diffusion and structural MRI modalities over time.

#### 2.2.5 Brain-Behavior validation across performance domains

The prior sections established data-driven evidence for a “best performing” configuration for NNHEmbed by using three partitions of the UKB and one specific cognitive outcome from the NNL with a UKB counterpart. We now quantify human performance validity of the NNHEmbed embeddings using additional cognitive scores from the Normative Neuroimaging Library (NNL) focusing on other aspects of healthy aging and cognition and two previously unused datasets: the Alzheimer’s Disease Neuroimaging Initiative (ADNI) encompassing the aging context (neurodegenerative disease participants were excluded) and the Parkinson’s Progression Markers Initiative (PPMI) centered on Parkinson’s disease populations. Each dataset provides a unique context to assess the robustness of the NNHEmbed-derived features in predicting cognitive and functional outcomes. Validation dataset demographics are in Table 8 and 9 with more detail in the methodology section along with cognitive/functional measure definitions.

**Table 8:** Baseline ADNI Population Characteristics.

| Characteristic | Female N = 152<sup>†</sup> | Male N = 99<sup>†</sup> |
| --- | --- | --- |
| AGE | 69 (65, 73) | 71 (67, 76) |
| PTEDUCAT | 16.00 (15.50, 18.00) | 18.00 (16.00, 20.00) |
| Race |  |  |
| non.white | 31 (20%) | 10 (10%) |
| white | 121 (80%) | 89 (90%) |
| MMSE | 29.00 (29.00, 30.00) | 29.00 (28.00, 30.00) |
| CDRSB | 0.00 (0.00, 0.00) | 0.00 (0.00, 0.50) |
| FAQ | 0.00 (0.00, 0.00) | 0.00 (0.00, 0.00) |
| Unknown | 4 | 2 |
| DTI_dti_FD_mean | 1.68 (1.19, 2.19) | 1.80 (1.42, 2.31) |
| rsfMRI_fcnxpro122_FD_mean | 0.12 (0.08, 0.16) | 0.11 (0.07, 0.16) |
| T1Hier_resnetGrade | 1.77 (1.47, 2.06) | 1.53 (1.24, 1.86) |
| snr | 10.3 (7.7, 13.8) | 8.8 (5.5, 12.1) |
| joinedDX |  |  |
| CN | 42 (28%) | 26 (26%) |
| MCI | 14 (9.3%) | 19 (19%) |
| SMC | 94 (63%) | 54 (55%) |
| Unknown | 2 | 0 |

<sup>†</sup>Median (Q1, Q3); n (%)

**Table 9:** Followup ADNI Population Characteristics.

| Characteristic | Female N = 52<sup>†</sup> | Male N = 41<sup>†</sup> |
| --- | --- | --- |
| **AGE** | 69 (66, 75) | 73 (68, 76) |
| **PTEDUCAT** | 18.00 (15.00, 18.00) | 18.00 (16.00, 19.00) |
| **Race** |  |  |
| non.white | 8 (15%) | 4 (9.8%) |
| white | 44 (85%) | 37 (90%) |
| **MMSE** | 30.00 (29.00, 30.00) | 29.00 (28.00, 30.00) |
| Unknown | 23 | 14 |
| **CDRSB** | 0.00 (0.00, 0.00) | 0.25 (0.00, 0.50) |
| Unknown | 23 | 13 |
| **FAQ** | 0.00 (0.00, 0.00) | 0.00 (0.00, 0.00) |
| Unknown | 24 | 13 |
| **DTI_dti_FD_mean** | 1.77 (1.20, 2.18) | 2.05 (1.56, 2.26) |
| **rsfMRI_fcnxpro122_FD_mean** | 0.13 (0.09, 0.15) | 0.11 (0.08, 0.17) |
| **T1Hier_resnetGrade** | 1.64 (1.44, 1.96) | 1.49 (1.26, 1.67) |
| **snr** | 12.4 (7.7, 14.4) | 9.0 (5.4, 11.4) |
| **joinedDX** |  |  |
| CN | 18 (35%) | 10 (24%) |
| MCI | 7 (13%) | 10 (24%) |
| SMC | 27 (52%) | 21 (51%) |

<sup>†</sup>Median (Q1, Q3); n (%)

The NNHEmbed embeddings derived from the UK Biobank were applied to each validation dataset using the same linear transformation learned during training. This ensured that the embeddings were directly comparable across datasets. We then conducted a series of cross-sectional and longitudinal regression analyses to assess the associations between these embeddings and relevant cognitive and functional measures. The same covariates were used across studies: brain volume, chronological age, biological sex and educational attainment (years). The individual cognitive or clinical measurements were collected into groups of variables with similar performance domains (e.g., memory, executive function, motor) to facilitate interpretation of the results. The cognitive variable categorizations are summarized in Figure 6. Results of the associations – mapped to Cohen’s f-squared effect sizes – are summarized in Figure 7. The results demonstrate that the NNHEmbed embeddings capture meaningful brain-behavior relationships across diverse populations and cognitive domains, underscoring their potential utility in clinical and research settings. Figure 8 shows a heatmap representing top features (imaging derived phenotypes, IDPs) for each key embedding and their associated cognitive domains. A significance threshold of *p* <= 0.001 was applied to identify robust associations (results are similar with a FDR-corrected threshold of *q* <= 0.05), with effect sizes interpreted using conventional benchmarks (small: *f* ^2^ = 0.02, medium: *f* ^2^ = 0.15, large: *f* ^2^ = 0.35).

**Figure 6.**
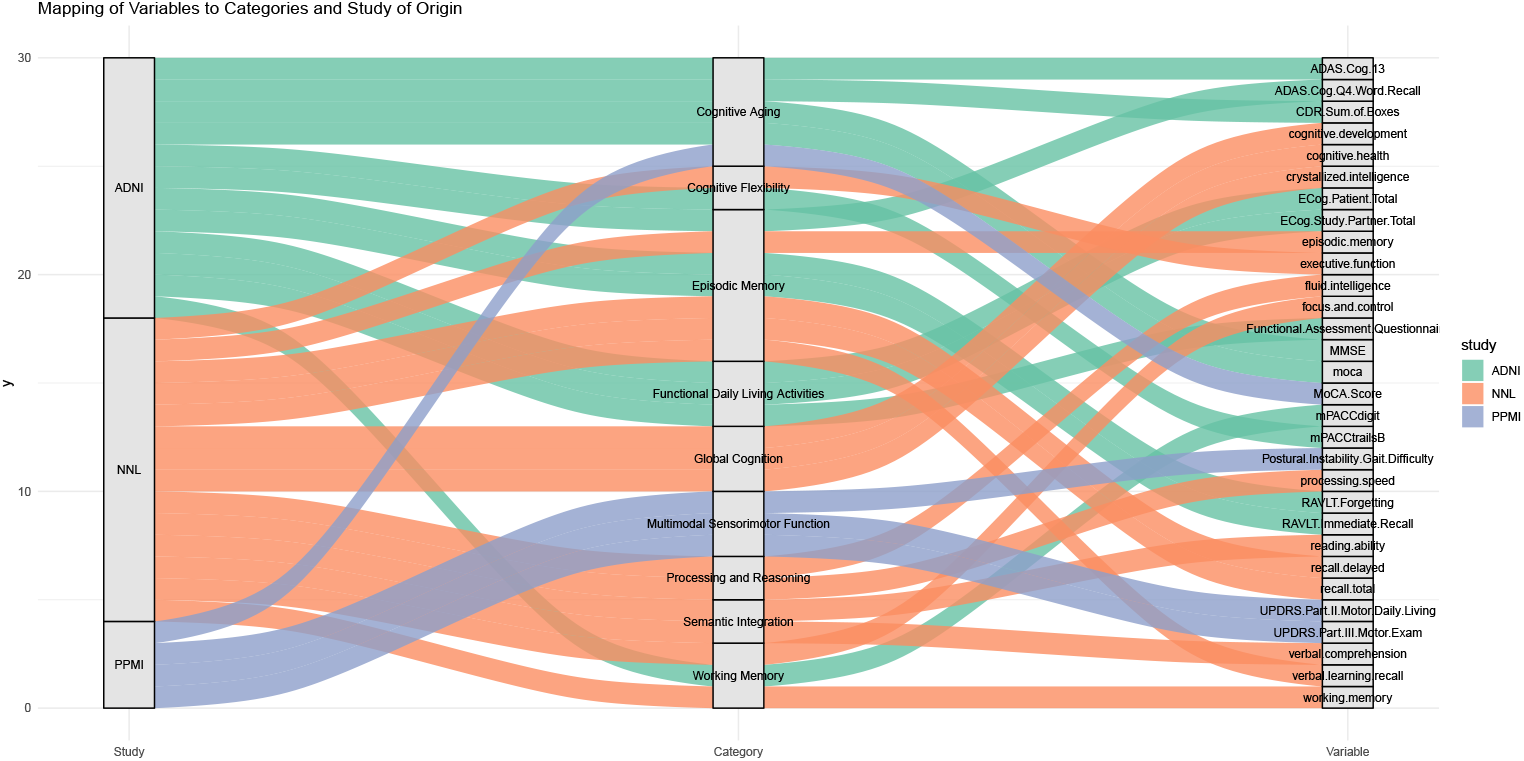
Cognitive Variables Categorized by Study and Measurement Type. Each cognitive variable from the NNL, ADNI, and PPMI studies is categorized by its respective study and measurement type. This categorization aids in interpreting the associations between brain imaging features and cognitive outcomes.

**Figure 7.**
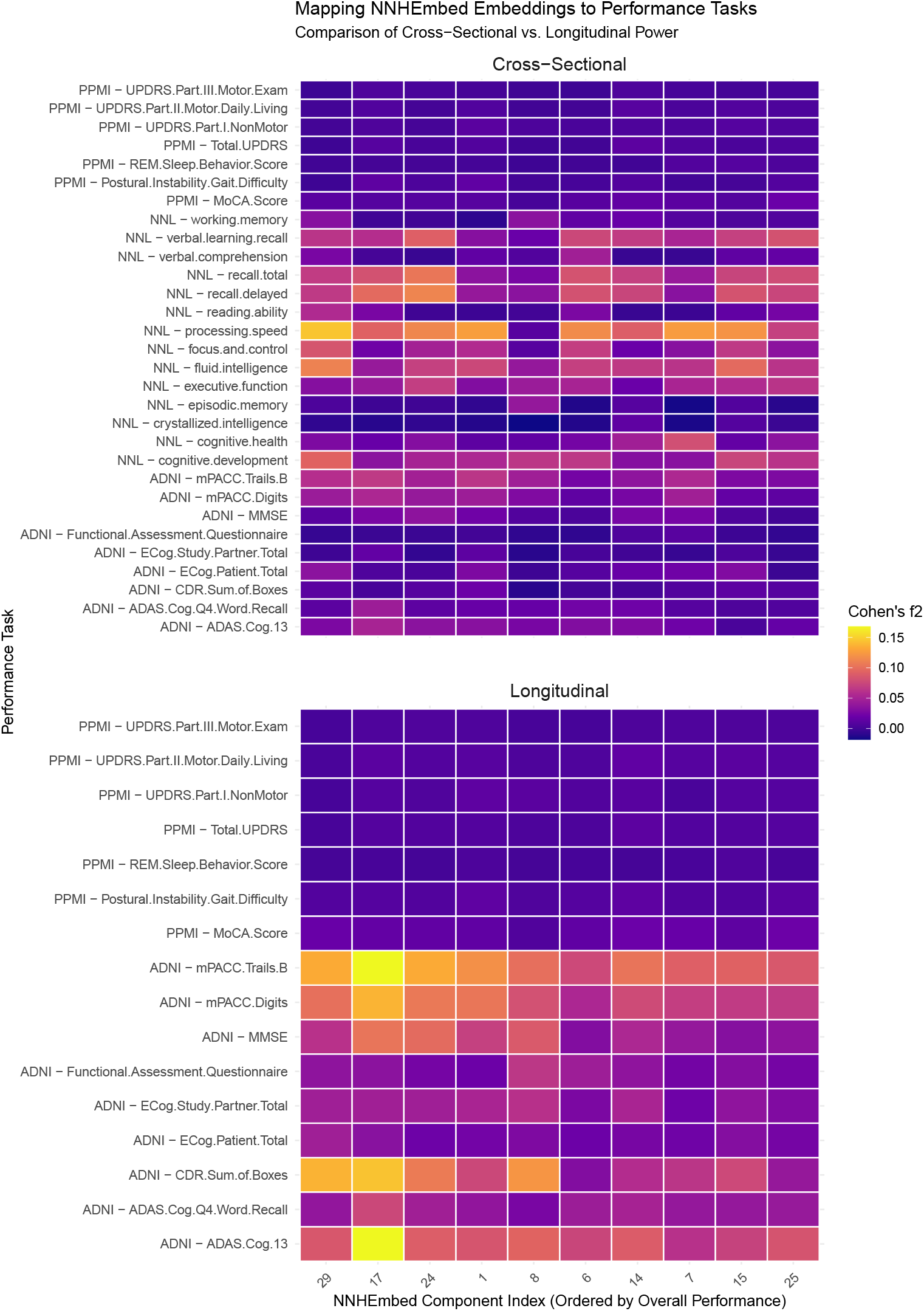
Heatmap of Top NNHEmbed Components Across All Performance Tasks. This heatmap visualizes the effect sizes (Cohen’s f-squared) of the top NNHEmbed components across various cognitive and clinical performance tasks from multiple datasets. Results show significant associations (FDR *q*-value <= 0.05). The color gradient indicates the strength of the association, with brighter colors representing higher effect sizes.

**Figure 8.**
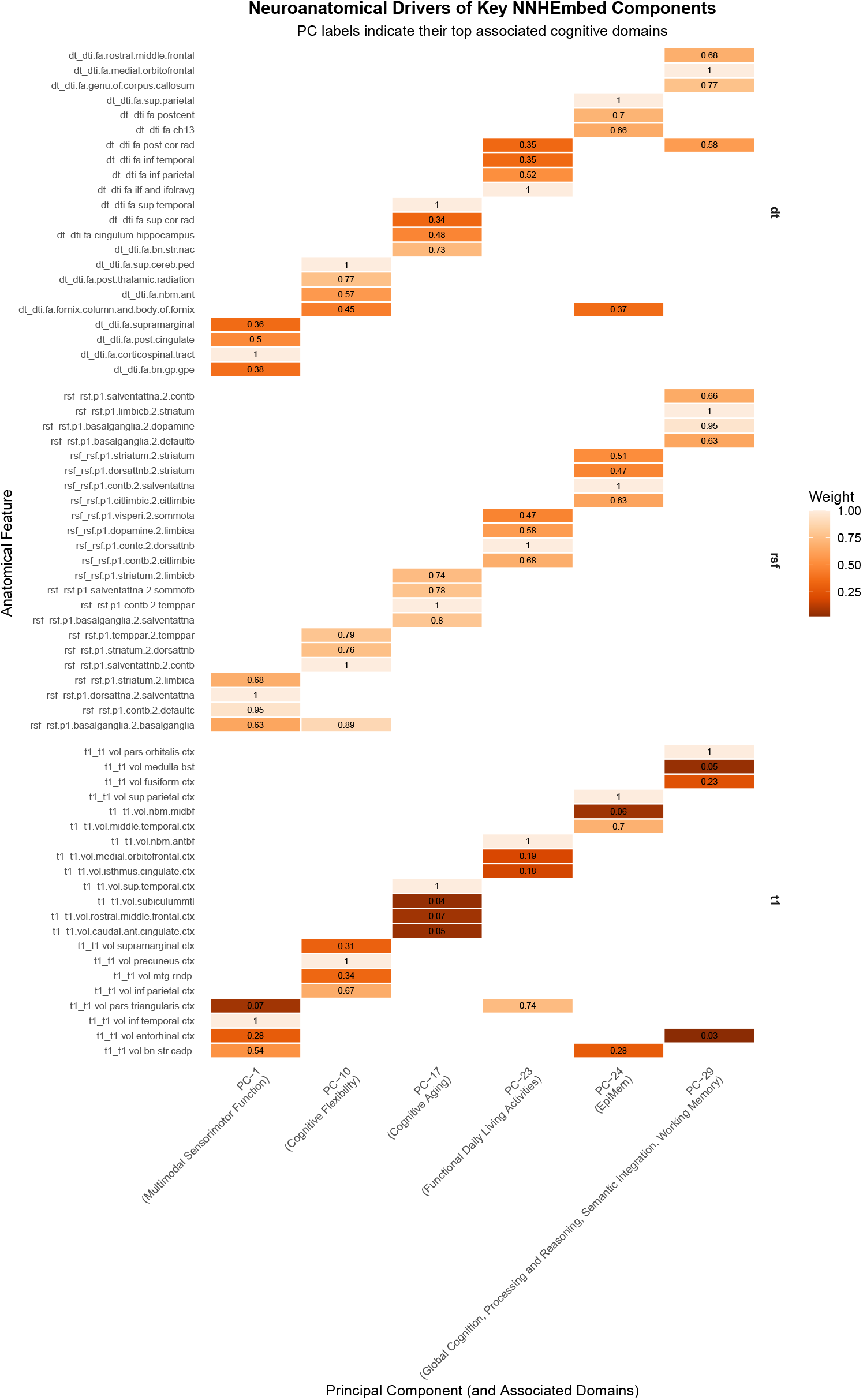
Heatmap of top feature weights for each key PC.

#### 2.2.6 Assignment of performance domains to embeddings

To link cognitive domains to each of the 31 NNHEmbed embeddings (i.e. where a component is the sMRI, dMRI and rsfMRI contribution acting as a whole) a maximum-only matching approach was used. This assigns each cognitive domain to the embedding with the strongest association based on Cohen’s *f* ^2^ effect sizes from cross-sectional data in NNL, PPMI and ADNI. This approach identifies the “champion” embedding for each domain but does not account for cases where multiple embeddings show similar strengths of association. The diversity of brain-to-behavior mappings therefore is reflective of the uniqueness of the underlying embedding and its emergent cognitive profile. A summary of the associations is shown in Figure 8. Below, we expand the shorthand names for the most relevant IDPs and detail the key neurobiolgical contributors to each of the top brain-behavior associations. We also comment on the potential mechanistic relevance of these data-driven results. Recall that these associations follow from the data-driven empirical analysis alone; thus, the interpretation is provisional and should be refined and/or validated via additional targeted studies.

##### 2.2.6.1 PC-1 → Multimodal Sensorimotor Function – Interpretation

A multimodal sensorimotor–limbic integration circuit: inferior temporal cortex and supramarginal gyrus provide higher-order perceptual inputs, while caudate, globus pallidus externus, and basal ganglia networks contribute action selection. Entorhinal cortex and posterior cingulate engage mnemonic integration, and corticospinal involvement anchors the component in embodied motor output. Supports visually grounded sensorimotor coordination and perceptual– motor integration.

###### Plausibility

Strong sensorimotor–basal ganglia motif with convergent temporal and cingulate contributions; multimodal white-matter/cortical patterns align with established perception–action pathways.

##### 2.2.6.2 PC-10 → Cognitive Flexibility – Interpretation

A cognitive flexibility and task-switching network: precuneus, inferior parietal lobule, and supramarginal gyrus form the classic frontoparietal hub for reorienting attention. Red nucleus and superior cerebellar peduncle contribute motor–cognitive coordination, while posterior thalamic radiation and fornix integrate thalamocortical and memory signaling. Basal nucleus of Meynert and salience→FPCN interactions support state shifting and attentional gating. Enables adaptive behavior under changing task demands.

###### Plausibility

Frontoparietal–salience–basal ganglia architecture strongly linked to cognitive flexibility; cerebellar–red-nucleus involvement consistent with motor–cognitive coupling.

##### 2.2.6.3 PC-17 → Cognitive Aging – Interpretation

A cognitive-aging vulnerability circuit: superior temporal cortex and temporoparietal networks reflect sensory-cognitive decline trajectories, while nucleus accumbens introduces motivational and reward sensitivity shifts common in aging. Cingulum-hippocampus and superior corona radiata emphasize memory and frontoparietal disconnection. Basal-ganglia→salience and salience→somatomotor transitions suggest progressive reduction in network segregation with age. Supports global cognitive slowing and reduced network specificity.

###### Plausibility

Multimodal evidence routinely links temporoparietal, basal forebrain, limbic, and frontoparietal disconnection with aging; pattern aligns with known age-related network alterations.

##### 2.2.6.4 PC-23 → Functional Daily Living Activities – Interpretation

A cholinergic–frontoparietal–visuomotor daily-function network: anterior basal nucleus of Meynert anchors cholinergic modulation, while pars triangularis and inferior parietal regions support language and praxis. Inferior temporal contributions reflect object recognition demands. White-matter pathways (IFOF, posterior corona radiata) connect perceptual and executive domains. Dopaminergic→limbic and frontoparietal→attention transitions indicate motivational and control components of real-world functioning. Supports everyday activity planning and goal-directed behavior.

###### Plausibility

Cholinergic basal forebrain + frontoparietal + IFOF patterns closely tied to functional daily living metrics; dopaminergic-limbic links are consistent with motivation dependencies.

##### 2.2.6.5 PC-24 → Episodic Memory – Interpretation

A temporal– parietal–prefrontal episodic memory system: superior parietal and middle temporal regions support multimodal encoding, while caudate and basal forebrain (Ch13) contribute strategic memory control and cholinergic enhancement. Fornix involvement identifies the canonical hippocampal output route. Frontoparietal→salience and limbic–striatal interactions modulate recall monitoring and memory salience. Supports episodic retrieval and associative binding.

###### Plausibility

Parietal–temporal + fornix ensemble is a hallmark episodic memory network; limbic and salience modulation well supported across imaging studies.

##### 2.2.6.6 PC-29 → Global Cognition, Processing and Reasoning, Semantic Integration, Working Memory – Interpretation

A general, high-order reasoning, semantic, and working-memory control network: pars orbitalis and medial orbitofrontal cortex provide semantic and valuation inputs; rostral middle frontal gyrus contributes executive planning. The genu of the corpus callosum and posterior corona radiata support interhemispheric and long-range integration. Limbic→striatal, basal-ganglia→dopaminergic, and salience→FPCN transitions reflect hierarchical cognitive control and motivation. Supports abstract reasoning, rule maintenance, and conceptual integration.

###### Plausibility

Prefrontal semantic/valuation regions combined with FPCN and dopaminergic–striatal pathways fit well with high-level cognition and working memory models.

#### 2.2.7 NNH health assessment at individual level

To demonstrate the clinical utility of the NNHEmbed features, we present a case study involving the normative profiling of an individual without clinical diagnosis of neurodegenerative disease from the ADNI cohort. This approach visualizes an individual’s brain metrics in relation to a large normative distribution, offering a quantitative method for tracking neurological status over time. This individual’s data was assessed over time: baseline and follow-up visit four years later. For each time point, a feature set—comprising demographic data (age, sex), cognition, and several NNHEmbed-derived embeddings (T1- and DTI-based PCs with targeted longitudinal and domain-specific interpretations)—was used to generate a normative summary report.

This longitudinal assessment (Figure 9) demonstrates how interpretable NNHEmbed features can be used to track an individual’s trajectory relative to a normative model. While not intended for diagnosis, this method provides an objective, data-driven profile that could aid clinicians in monitoring exposure-related, adaptive or compensatory changes and assessing an individual’s deviation from an expected trajectory. In this case, changes over the visits are demonstrated that may also relate to changes in cognition, here represented by the ADAS13 score (higher is more impairment).

**Figure 9.**
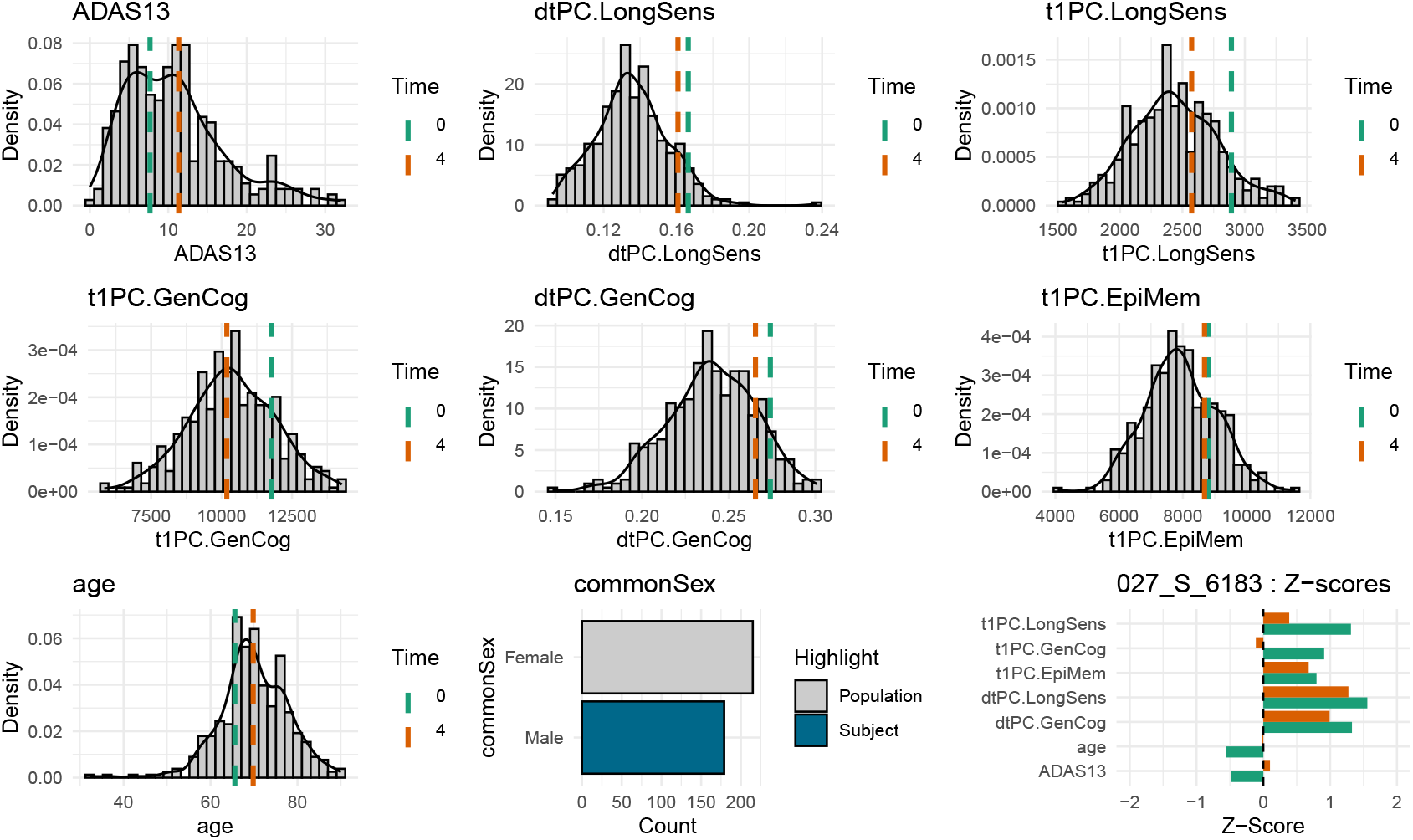
Normative summary plots for an ADNI control subject at baseline and follow-up. The subject data is compared to a normative distribution where we visualize how the subject values shift over time. The subjects ADAS13 (cognitive impairment) score increases while the longitudinally and cognitively specific sMRI, dMRI embeddings show evidence of atrophy.

From a dimensional perspective, the subject’s T1-weighted longitudinal sensitivity component (t1PC.LongSens) and T1 and diffusion-derived general cognition sensitivity component (*PC.GenCog) both show gradual declines across the available observations. These trajectories suggest subtle but consistent movement away from normative expectations, consistent with age-related decline. This aligns with the modest but detectable increase in ADAS13 score (7.67 → 11.33). The concordance between behavioral and imaging-derived PC trajectories highlights the potential sensitivity of the NNHEmbed framework to clinically relevant change.

Overall, these PC-derived measures contextualize the subject’s aging profile within a broader normative framework. Although still within a range compatible with healthy aging, the pattern of decline across longitudinal sensitivity and cognitive components suggests that this individual may be at elevated risk for future cognitive impairment. Additional prospective application will be needed to determine the prognostic utility of these multi-modal embeddings.

#### 2.2.8 NNH assessment at population level

To demonstrate the clinical utility of the NNHEmbed features, we first presented a single-subject case study from the ADNI cohort (Figure 9). That analysis illustrated how individualized brain–behavior trajectories can be contextualized within a large, multi-modal normative model, revealing subtle deviations in longitudinal sensitivity and episodic-memory–related components consistent with mild age-related cognitive change.

We next extended this approach to characterize group-level normative trajectories across major diagnostic categories in ADNI: cognitively normal (CN), mild cognitive impairment (MCI), and Alzheimer’s disease (AD). This analysis generates pseudo-average subjects by computing the mean feature trajectory over three follow-up intervals (baseline, year 1, and year 2) for each diagnosis, and compares these averaged values against the normative baseline derived from healthy control participants.

Each “pseudo-subject” represents the typical pattern of longitudinal change within that group, allowing direct visualization of how canonical NNHEmbed-derived features deviate from healthy norms. All models included demographic variables (age, sex), motion-adjusted diffusion components, and domain-specific principal components capturing longitudinal sensitivity, processing speed, and episodic memory. The resulting normative reports (Figures 10–12) summarize both the direction and magnitude of deviations relative to the control population.

**Figure 10.**
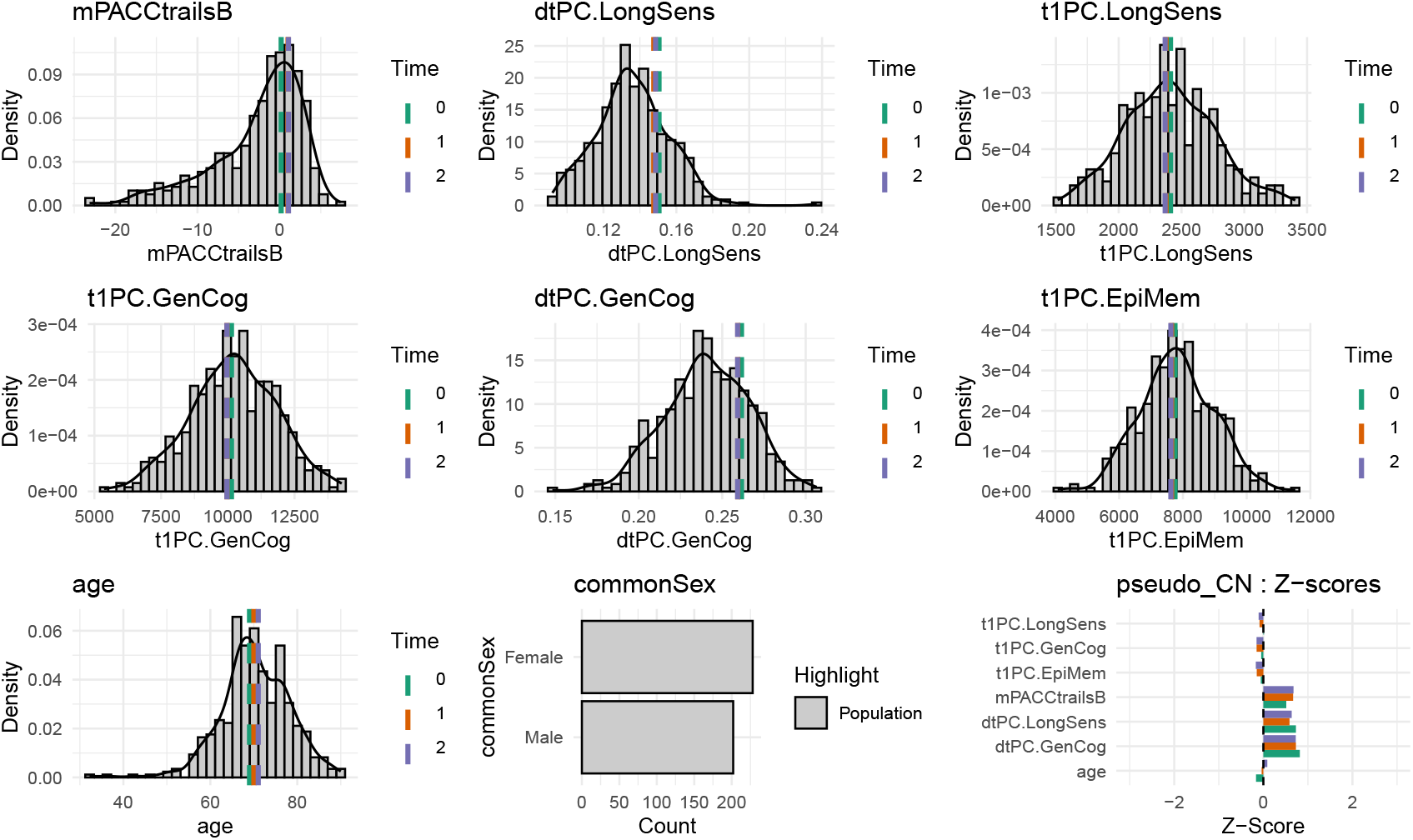
Normative summary for the cognitively normal (CN) group. Mean trajectories across three visits (baseline, year 1, year 2) show minimal deviation from normative expectations, with stable patterns in longitudinal sensitivity and cognitive components. See the barplots at lower right for the quickest summary visualization.

**Figure 11.**
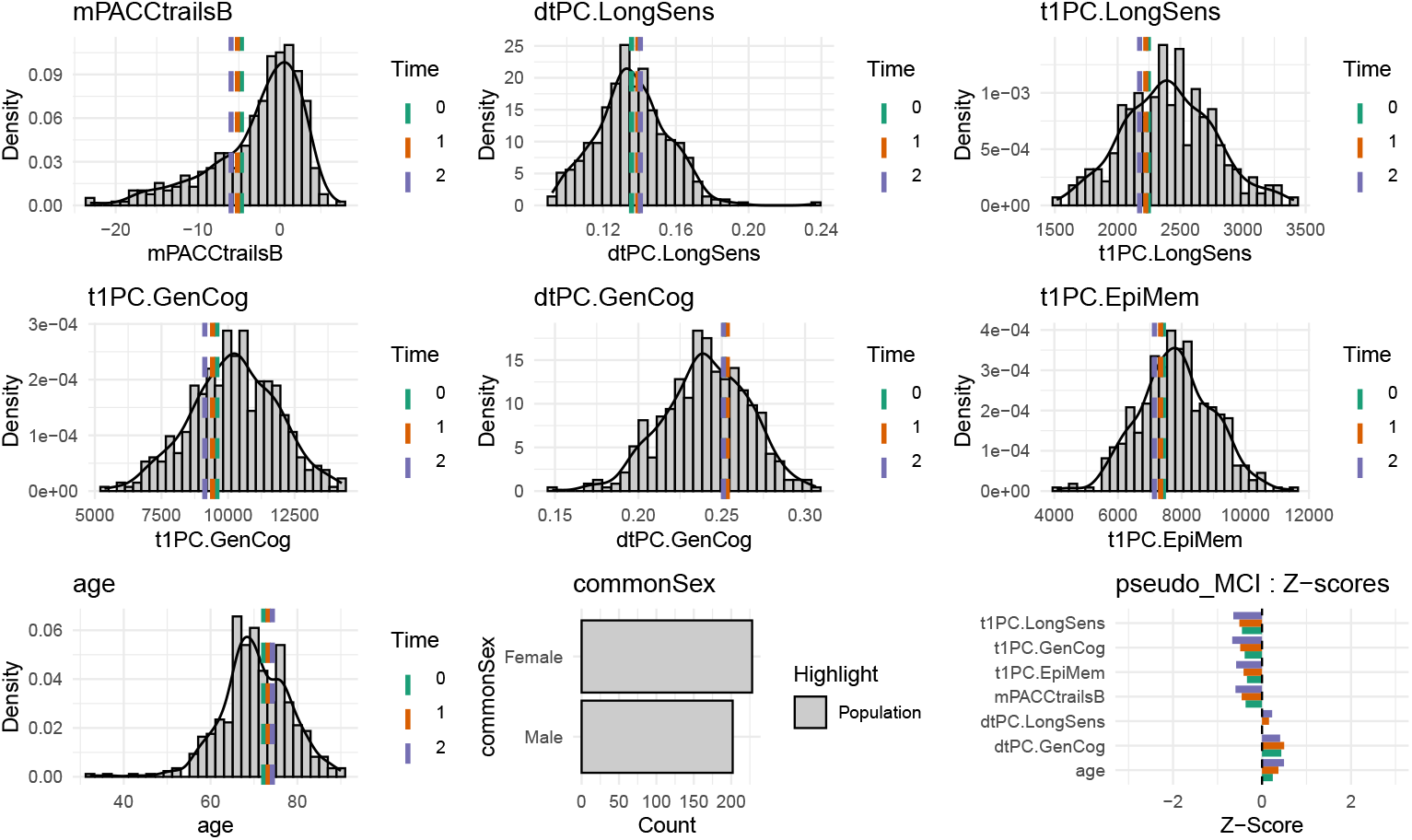
Normative summary for the mild cognitive impairment (MCI) group. Mean trajectories across three visits (baseline, year 1, year 2) show moderate deviation from normative expectations, with somewhat reduced patterns across summary components. See the barplots at lower right for the quickest summary visualization.

**Figure 12.**
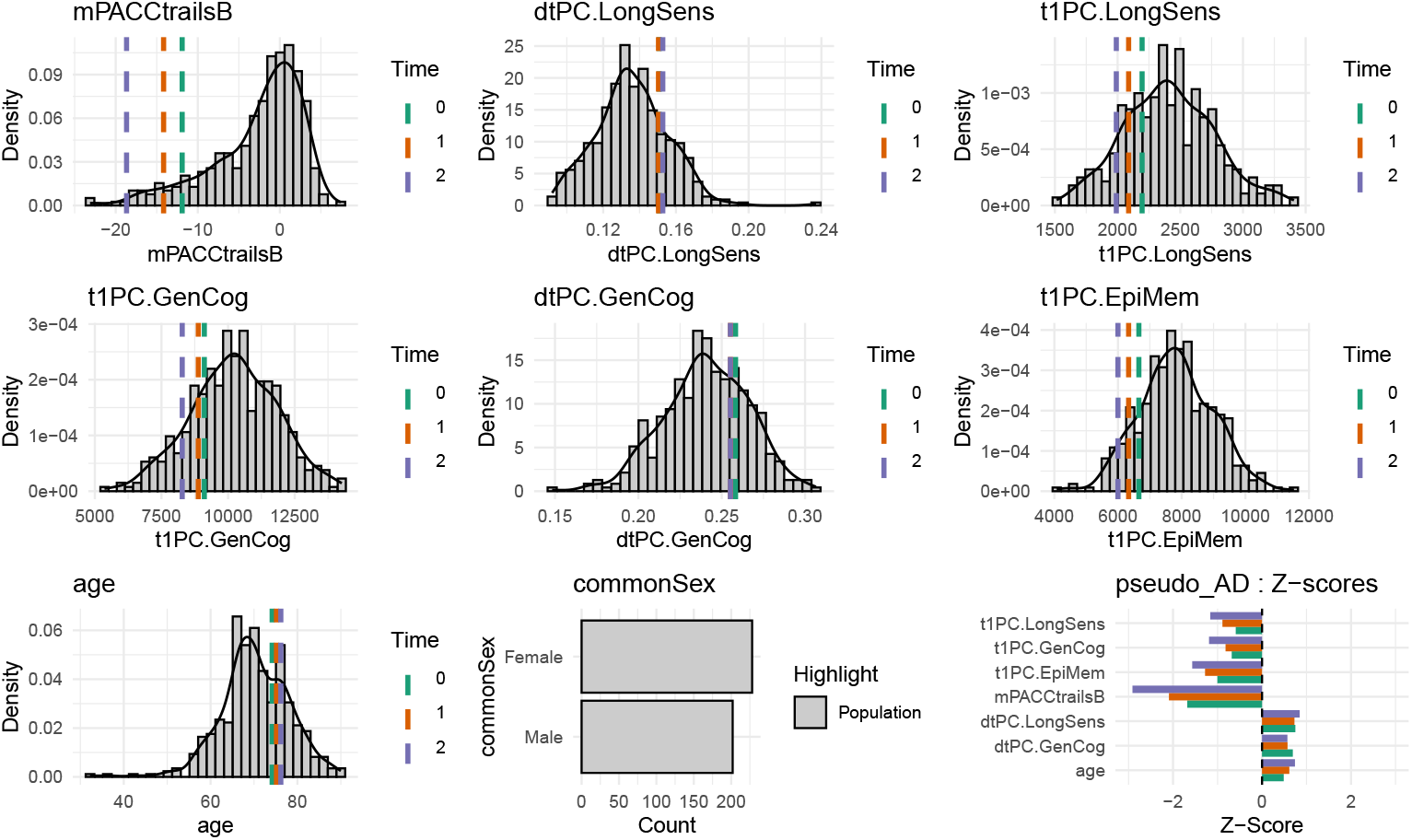
Normative summary for the AD group. Mean trajectories across three visits (baseline, year 1, year 2) show pronounced deviation from normative expectations.

Across groups, progressive divergence from the normative model is evident. Cognitively normal (CN) individuals show stable feature trajectories clustered near the mean of the normative distribution. MCI participants exhibit small but systematic shifts in the components, consistent with mild decline in network integrity and cortical volume associated with early-stage neurodegeneration. By contrast, AD participants demonstrate broad deviations across nearly all feature domains, including pronounced reductions in scores, indicating coordinated deterioration across structural and diffusion-based representations of brain health.

This analysis confirms that the NNHEmbed features capture meaningful variation across clinical stages while maintaining direct interpretability at the subject level. The longitudinal summaries can thus serve both as individual monitoring tools and as quantitative phenotypes for studying group-level trajectories in neurodegenerative disease progression.

Together, these visual and quantitative analyses confirm that the NNHEmbed framework yields biologically interpretable representations that scale from individual to population-level inference. They provide a practical bridge between deep feature learning and normative modeling, highlighting how latent multimodal embeddings can be used to track and contextualize neurodegenerative trajectories.

## 3 Discussion

This study systematically evaluated the NNHEmbed framework for learning joint representations from multi-modality MRI and assessed utility for brain-behavior mapping across diverse cohorts and longitudinal settings. We demonstrated that NNHEmbed, utilizing different cost functions, can effectively integrate information from sMRI, rsfMRI, and dMRI to generate meaningful low-dimensional representations that are sensitive to cognitive function, clinical status, and longitudinal change. In particular, the best performing model – identified by data-driven evaluations – revealed neuroscientifically interpretable systems in the brain. While these systems are indeed complex and multi-faceted, they align well, overall, with established knowledge of brain organization and function.

Our unsupervised evaluation on the UK Biobank provided a critical foundation, allowing selection of robust NNHEmbed parameter sets based on cross-study prediction power, representation quality and longitudinal sensitivity, independent of the specific behavioral outcomes tested later. This multi-phase approach enhances the reliability and generalizability of the findings. The comparison between cost functions (e.g. regression (Reg), normalized correlation (NC), log-cosh, and Absolute Canonical Covariance (ACC)) revealed that the latter and the closely related NC provided overall best results. In particular, NNHEmbed-ACC with ICA as mixing method and orthogonality constraints produced the best result overall. However, we note that the choice between energy functions should be guided by the specific research question at hand and that these results may not generalize to new problems. Notably, the baseline methods (both PCA and CCA based) did not score well enough in our evaluation metrics in order to breach the top 20 rankings, underscoring the advantage of NNHEmbed multi-view learning approaches for complex neuroimaging data.

The successful application of UKB-derived bases to the NNL, ADNI, and PPMI cohorts underscores the potential for leveraging large population datasets to create generalizable normative brain representations in a multi-view context. The NNHEmbed embeddings captured significant variance related to cognitive performance and aging-related clinical presentation. Crucially, the multi-modal representations in NNHEmbed components demonstrated the capacity to map to specific cognitive domains without direct supervision by those domains. This may arise from the large-scale training data available in the UKB and the multi-modal nature of the data, which together enable the model to learn rich, biologically relevant patterns. Lastly, the demonstration of individual profiling against normative distributions illustrates a practical application of these NNHEmbed bases for tracking neurological health, potentially identifying response to experience or to intervention based on multi-modal brain signatures.

### Limitations and Future Directions

The multivariate nature of the derived embeddings, by necessity, loses some degree of granularity in comparison to traditional focal analyses of brain and behavior links as in single-subject lesion studies. This is perhaps a necessary compromise when analyzing data from high *n* healthy participant cohorts. Noise is further amplified by the imperfect nature of the psychometric and clinical measurements used here which are subject to age effects, learning effects and imperfect unimodal specificity. While standard cognitive tests were used, they represent only a fraction of human behavior. Additionallly, this analysis, while comprehensive, relies on aggregated metrics. Future work might dissect successes at a finer-grained level by investigating, for instance, *which specific PCs* are driving the predictions for the cross-study outcomes. Furthermore, the high-level nature of multivariate MRI-derived features suggests that additional data enabling a deeper biological interpretation would add value; e.g. correlating the top-performing PCs with known maps of neuronal cell types or gene expression profiles would be a crucial next step.

Additionally, the specific feature extraction and preprocessing steps influence the final representations. Harmonization across different scanners and cohorts remains a challenge, although using a fixed UKB-derived basis provides some robustness as demonstrated by cross-cohort prediction results. Finally, interpreting the precise neurobiological meaning of complex, multivariate components requires careful follow-up investigation beyond the scope of this initial evaluation.

## 4 Conclusion

In conclusion, this comprehensive evaluation successfully identifies an optimal end-to-end analysis strategy for converting high-dimensional M3RI into a panel that is appropriate for representing neurological health. Focusing, for example, only on the embeddings that are annotated with known relationships to the performance domains shown here (e.g. working memory, cognitive flexibility, episodic memory) would provide a compact and interpretable and accessible visual representation for future studies of aging, exposure, performance and/or risk. Additionally, the generated features are exceptionally potent for capturing longitudinal change, making them a powerful tool for studies of training or experience. However, we acknowledge that such data is relatively rarely available in non-clinical populations and that other broadly known technical challenges to longitudinal data collection remain.

This work is grounded in a biologically principled view of brain organization, wherein structure and function are deeply intertwined across spatial and temporal scales (Passingham, Stephan, and Kötter 2002; Mountcastle 1997). We align with emerging frameworks that conceptualize brain health not as binary but as a continuum, with pathology representing deviations from normative developmental and aging trajectories (Gaser, Kalc, and Cole 2024; Reicher et al. 2025). In this context, multi-view learning offers a principled strategy to capture and interpret M3RI enabling pattern identification that is relevant in normal populations and, potentially, clinical populations. Therefore, the results of this work will provide a foundation for future studies that seek to understand how multi-modal brain features relate to not only cognition and aging but also other contexts such as elevated performance, occupational exposures and novel experience across diverse populations.

## 5 Methods

### 5.1 Datasets

#### UK Biobank (UKB)

Primary dataset for training and parameter selection. We utilized M3RI data (T1-weighted sMRI, rsfMRI, dMRI) from N=21,300 participants selected from the larger imaging cohort (Miller et al. 2016) based on data availability and quality control. Table 4, 5 and 6 detail the number of subjects used for each component of the analysis (cross-sectional train, cross-sectional test and longitudinal validation). The UKB cohort is predominantly middle-aged to older adults (age range 44-82 years, mean age ~64 years).

#### Normative Neuroimaging Library (NNL)

A healthy participant dataset comprising M3RI data from adults across a wide age range (n=164). This dataset includes comprehensive cognitive assessments covering standard cognitive domains assessed in the NIH toolbox. NNL served as the primary validation set for cognitive associations in a healthy population. See Table 7 for demographic details.

#### Alzheimer’s Disease Neuroimaging Initiative (ADNI)

Used for validation in the context of aging and risk for Alzheimer’s disease (“Alzheimers Disease Neuroimaging Initiative (ADNI) Database,” n.d.). We included M3RI data and cognitive/functional scores (e.g., ADAS-Cog, CDR-SB, FAQ - see definitions below) from ADNI 1/GO/2/3 phases, encompassing healthy controls or subjective memory concern defined as MMSE ≥ 24 & DX ≠ AD. Both cross-sectional and longitudinal data (up to 5+ years) were analyzed. See Table 8 and 9 for demographic details.

#### Parkinson’s Progression Markers Initiative (PPMI)

Used for validation in the context of aging and aging-related motor disorders including Parkinson’s disease (“Parkinsons Progression Markers Initiative (PPMI) Database,” n.d.). We included M3RI data and cognitive/clinical scores (e.g., MoCA, MDS-UPDRS) from PPMI. Both cross-sectional and longitudinal data were utilized. See Table 10 for demographic details.

**Table 10:** Baseline PPMI Population Characteristics.

| Characteristic | Female N = 455<sup>†</sup> | Male N = 615<sup>†</sup> |
| --- | --- | --- |
| **age_BL** | 66 (62, 71) | 67 (62, 72) |
| **EDUCYRS** | 16.0 (14.0, 18.0) | 16.0 (15.0, 18.0) |
| Unknown | 6 | 7 |
| **Diagnosis** |  |  |
| CN | 25 (5.5%) | 35 (5.7%) |
| PDGBA | 7 (1.5%) | 6 (1.0%) |
| PDLRRK2 | 10 (2.2%) | 4 (0.7%) |
| PDPRKN | 0 (0%) | 4 (0.7%) |
| PDSNCA | 1 (0.2%) | 1 (0.2%) |
| PDSporadic | 134 (29%) | 251 (41%) |
| ProdromalGBA | 5 (1.1%) | 3 (0.5%) |
| ProdromalLRRK2 | 17 (3.7%) | 8 (1.3%) |
| ProdromalSporadic | 256 (56%) | 301 (49%) |
| SWEDDSporadic | 0 (0%) | 2 (0.3%) |
| **moca** | 28.00 (26.00, 29.00) | 27.00 (25.00, 29.00) |
| Unknown | 9 | 10 |
| **updrs_totscore** | 15 (7, 31) | 19 (9, 33) |
| Unknown | 23 | 29 |
| **pigd** | 0.00 (0.00, 0.20) | 0.00 (0.00, 0.20) |
| Unknown | 13 | 18 |
| **DTI_dti_FD_mean** | 1.58 (1.21, 2.17) | 1.60 (1.25, 2.33) |
| **rsfMRI_fcnxpro122_FD_mean** | 0.16 (0.12, 0.21) | 0.16 (0.12, 0.22) |
| **T1Hier_resnetGrade** | 1.96 (1.59, 2.29) | 1.74 (1.36, 2.13) |
| **snr** | 11.7 (8.8, 14.6) | 10.9 (8.0, 13.6) |

<sup>†</sup>Median (Q1, Q3); n (%)

### 5.2 ANTsPyMM-derived Imaging Phenotypes (IDPs)

All M3RI data underwent standardized preprocessing using ANTs tools along with their pythonic wrapping (Brian B. Avants et al. 2011) and ANTsR (Brian B. Avants et al. 2021). ANTsPyMM is a Python-based toolkit in the ANTsX ecosystem focused on reproducible computation for multi-modality MRI processing and IDP extraction, supporting modalities such as T1-weighted structural MRI, diffusion MRI, resting-state fMRI, arterial spin labeling (ASL), and metabolic imaging. oai_citation:0‡GitHub Below is a summary of key types of IDPs derived by ANTsPyMM, their neurobiological interpretation, and typical usage in brain-behavior mapping.

#### 5.2.1 Structural / T1-weighted MRI IDPs

##### Cortical and subcortical morphometry

Measures include cortical thickness, surface area, volume, and subcortical volumes (e.g. hippocampus, basal forebrain, cerebellum). These are computed after brain extraction, tissue segmentation (CSF, gray matter, white matter, deep nuclei), and parcellation into anatomical regions. oai_citation:1‡GitHub

- *Interpretation*: These IDPs quantify macroscopic brain structure, allowing investigation of atrophy, regional volumetric change, and structural correlates of cognition or disease.
- *Typical use*: As predictors or covariates in brain–behavior models, or as morphological anchors when integrating with diffusion or functional data.

##### Super-resolution interpolation / registration to template space

ANTsPyMM may apply super-resolution (SR) steps or precise registration of T1 images to a common template space (e.g. 0.5 mm isotropic) prior to quantification. oai_citation:2‡ResearchGate

- *Interpretation*: Improves spatial fidelity and inter-subject alignment, reducing partial-volume artifacts in morphometric measures.
- *Typical use*: Ensures consistent regional labeling across subjects and modalities.

#### 5.2.2 Diffusion MRI (dMRI) / Microstructure IDPs

##### Fractional Anisotropy (FA), Mean Diffusivity (MD), and related metrics

After motion correction, registration to T1 space, and diffusion model fitting (e.g. via DIPY), ANTsPyMM yields regional (atlas-based) summary statistics of FA, MD, and (potentially) other diffusion metrics (e.g. radial or axial diffusivity). oai_citation:3‡GitHub

- *Interpretation*: FA and MD characterize white matter microstructural integrity (e.g. degree of directional water diffusion, cellular barriers).
- *Typical use*: As correlates of connectivity, aging, pathology, or predictors of cognitive performance.

##### Tractography-based connectivity summaries

Dense tractography and connectivity matrices (e.g. streamline counts or weights between parcels) may also be output, along with regional fiber metrics.

- *Interpretation*: Provides a structural network view linking brain regions by estimated white-matter paths.
- *Typical use*: Graph-theoretic modeling, multimodal fusion with functional networks.

#### 5.2.3 Resting-State fMRI (rsfMRI) / Functional IDPs

##### Temporal statistics & motion metrics

Outputs include measures such as framewise displacement (FD), DVARS, temporal standard deviation (tSTD), signal-to-noise ratio (SNR), and motion-derived nuisance metrics (e.g. CompCor). oai_citation:5‡GitHub

- *Interpretation*: These quantify data quality, physiological noise, and signal stability, which are essential for later connectivity analysis.
- *Typical use*: As quality control covariates, screening, or correction variables.

##### Functional connectivity matrices (region-to-region correlations)

After preprocessing (motion correction, denoising, filtering), time series are averaged within parcels (e.g. via homotopic parcellation or Yeo parcellations) and Pearson (or similar) correlation or connectivity strength is computed between each region pair. oai_citation:6‡GitHub

- *Interpretation*: Represents synchronized activity across brain regions, reflecting functional networks (default mode, attention, visual, etc.).
- *Typical use*: As features in brain–behavior mapping, or functional network modes in multivariate models.

##### Derived metrics (e.g. fALFF, mPerAF, ALFF variants)

ANTsPyMM may compute amplitude-based metrics like fractional ALFF (fALFF), modified ALFF, or mean Percent Amplitude of Fluctuation (mPerAF) to quantify regional spontaneous activity amplitude. oai_citation:7‡ResearchGate

- *Interpretation*: Captures local fluctuation magnitude or baseline spontaneous activity variation.
- *Typical use*: Regional brain physiology marker, or as contrast features in functional regression models.

#### 5.2.4 Usage Notes & Considerations

##### Quality control & automated QC filters

As ANTsPyMM’s primary goal is robust IDP generation, it integrates an automated T1-weighted image quality reviewer (e.g. resnetGrader) to flag low-quality scans before subsequent processing. oai_citation:10‡ResearchGate

- Poor-quality T1 images may abort the pipeline or be excluded, because accurate anatomical labels underpin downstream dMRI and rsfMRI alignment.
- Quality control meta-data is included in the output for user review e.g. motion metrics, SNR, etc.

##### Neuroanatomical labeling & coordinate systems

The outputs are linked to standard parcellations (e.g. homotopic parcellation, DKT, JHU white-matter atlas) to facilitate interpretability and cross-subject alignment. oai_citation:11‡ResearchGate

##### Modality missingness

ANTsPyMM is designed to be modular: not all modalities are strictly required for a subject; missing modalities can be handled gracefully in downstream analyses. oai_citation:12‡ResearchGate

##### Interpretation caution for tractography

While tractography outputs are provided, we note that performance evaluation for dense tractography is ongoing and recommend cautious use (e.g. preferring summary connectivity measures over voxel-level streamlines). oai_citation:13‡ResearchGate

##### Open source & reproducibility

ANTsPyMM is open-source, and its processing steps are transparent, facilitating reproducibility and extensibility in large-scale, cross-study datasets. Docker files enabling containerization are also provided. oai_citation:14‡GitHub

Processed voxel-wise maps (or regional summaries, e.g., based on standard atlases) from each modality (e.g., GM density, cortical thickness, ALFF, FA) were vectorized per subject and compiled into modality-specific matrices 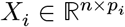, ensuring subject alignment across matrices and enabling multi-view learning. Each matrix was normalized by the sum of its eigenvalue spectrum to balance variance contributions across modalities and whitened to standardize feature scales. Subjects with missing modalities were excluded from the NNHEmbed training set to ensure complete data for multi-view integration.

### 5.3 NNHEmbed Framework

Normative Neurological Health Embedding (NNHEmbed) is a flexible framework for deriving low-dimensional, interpretable embeddings from highdimensional, multi-modal data. It extends methods like PCA and ICA by leveraging inter-modality relationships through a reciprocal modeling approach. Implemented in the ANTsR package, NNHEmbed uncovers shared latent structures in datasets such as neuroimaging or multi-omics data (Brian B. Avants et al. 2021). Let there be *Z* modalities (views), such as cortical thickness, white matter diffusion metrics, or functional connectivity. Each modality is represented as a data matrix

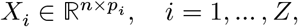

where *n* is the number of samples (subjects) and *p*_*i*_ is the number of features in modality *i*. NNHEmbed assumes a **shared latent embedding** *U* ∈ ℝ^*n*×*k*^, with *k* ≪ min_*i*_ *p*_*i*_, capturing common variation across all modalities. Each modality is linearly mapped to this shared embedding through a matrix

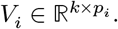

The columns of *U* represent subject-level latent scores, while the rows of *V*_*i*_ represent multivariate loadings (e.g., spatial maps for brain regions or connectivity measures). The embedding *U*_≠*i*_, approximating the true *U*, is derived from the concatenated set of 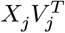 for all *j* ≠ *i*, using dimensionality reduction techniques like PCA, ICA, or SVD.

#### Optimization Objective

The NNHEmbed optimization problem is defined as:

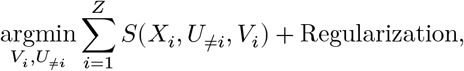

where:

- *i* indexes modalities from 1 to *Z*.
- *S* is a similarity function measuring the alignment between modalities.
- 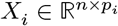 is the data matrix for modality *i*.
- *U*_≠*i*_ ∈ ℝ^*n*×*k*^ is the low-dimensional embedding derived from all modalities except *X*_*i*_, formed by applying PCA, ICA, SVD, or averaging over 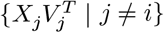.
- 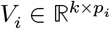 is the latent feature matrix for modality *i*, analogous to regression coefficients or loadings.
- Regularization is a sum of terms that impose structure on *V*_*i*_ (e.g., sparsity, smoothness, orthogonality) or may include domain knowledge priors.

#### 5.3.1 Energy Functions

The data fidelity term *S*(*X*_*i*_, *U*_≠*i*_, *V*_*i*_) supports multiple objectives, each emphasizing different properties of the latent space. Below, we detail each energy function, its interpretation, strengths and limitations.

##### 5.3.1.1 Regression / Reconstruction Error Definition

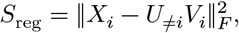

or its centered/normalized variants.

**Interpretation:** Encourages *U*_≠*i*_*V*_*i*_ to reconstruct *X*_*i*_, capturing maximum variance within modality *i*.

**Strengths:** - Convex in *V*_*i*_, simplifying optimization. - Prioritizes within-modality data fidelity, ensuring high variance capture.

**Limitations:** - Emphasizes within-modality variance, potentially neglecting cross-modality alignment. - Sensitive to noise and outliers, which may dominate the Frobenius norm.

##### 5.3.1.2 Normalized Correlation / Procrustes Correlation Definition

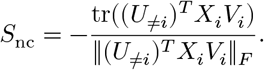

**Interpretation:** Maximizes correlation (up to rotation/scale) between *X*_*i*_*V*_*i*_ and *U*_≠*i*_, similar to canonical correlation analysis; normalized by the Frobenius norm.

**Strengths:** - Scale-invariant, focusing on geometric alignment of subspaces. - Enhances cross-modality similarity, supporting multimodal integration.

**Limitations:** - Non-convex, sensitive to initialization and optimization challenges. - May prioritize alignment over reconstruction accuracy.

##### 5.3.1.3 Absolute Canonical Covariance Definition

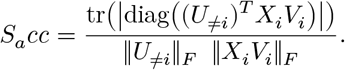

**Interpretation:**

Measures *dimensionwise canonical alignment* between the joint latent representation *U*_≠*i*_ and the modality-specific projection *X*_*i*_*V*_*i*_. Taking the absolute diagonal ensures robustness to sign ambiguity, while Frobenius normalization yields a scale-invariant alignment score.

**Strengths:**

- Captures **direct, axis-specific correspondence** between latent factors.
- **Robust to sign flips**, preserving meaningful canonical relationships.
- **Scale-invariant**, enabling cross-modality comparison of alignment strength.

**Limitations:**

- Ignores **off-diagonal cross-factor alignment**, which may carry additional structure.
- Sensitive when either denominator is small.
- Prioritizes alignment quality over reconstruction fidelity.

##### 5.3.1.4 Independence / non-Gaussianity Definition (logcosh example)

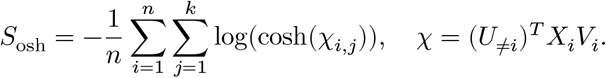

**Interpretation:** As with all of these metrics, this is optimized with respect to the *V*_*i*_ and, as such, promotes non-Gaussianity within the low-dimensional shared representation. The logcosh function approximates negentropy, encouraging sparse or independent latent factors, useful for separating distinct neural processes (e.g., motor vs. memory signals).

**Strengths:** - Captures sparse or independent factors, aligning with biological assumptions. - Robust to Gaussian noise due to non-Gaussianity focus.

**Limitations:** - Sensitive to non-linearity choice (e.g., logcosh) and data scaling. - May overfit to non-Gaussian artifacts without careful tuning.

##### 5.1.3.5 Domain Knowledge Priors Definition

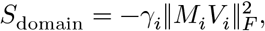

where 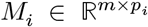 encodes prior information (e.g., anatomical or cognitive domain mappings).

**Interpretation:** Encourages *V*_*i*_ to align with prior knowledge by maximizing the energy of *M*_*i*_*V*_*i*_. In neuroimaging, *M*_*i*_ may represent cognitive domains, ensuring learned factors respect biological priors.

**Strengths:** - Integrates priors (e.g., anatomical atlases) for interpretable solutions. - Flexible: *M*_*i*_ can be sparse or derived from prior studies. - Supports hypothesis-driven research in neuroscience.

**Limitations:** - Requires accurate priors; restrictive *M*_*i*_ may bias results. - Performance depends on the tuning parameter *γ*_*i*_.

This quadratic regularizer penalizes deviation from the domain-consistent sub-space, guiding optimization toward solutions that reflect the prior structure.

#### 5.3.2 Summary of Energy Functions

Each energy function targets a distinct property:

- **Regression**: Prioritizes within-modality variance for data reconstruction.
- **Normalized Correlation**: Enhances cross-modality subspace alignment.
- **Absolute Canonical Covariance**: Ensures robust canonical relationships.
- **ICA**: Disentangles independent neural signals.
- **Domain Alignment/Domain Knowledge Priors**: Integrates priors to influence solutions toward interpretability.

#### 5.3.3 Regularization

Regularization is applied to the *V*_*i*_ matrices to impose structure (e.g., sparsity).

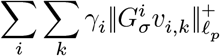

where *v*_*i,k*_ is the *k*-th feature vector in *V*_*i*_, *G*^*i*^ is a sparse regularization matrix, and *γ*_*i*_ is a scalar weight. The norm 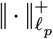is a positive-part *ℓ*_*p*_ norm, promoting sparsity or smoothness as needed where typically we use *ℓ*_1_. This regularization is invoke when orthogonality constraints are set to “none”.

Orthogonality constraints on the feature space encourage distinct feature representations across the full basis. When these are invoked (denoted in Table 1 as parameters with “orth”), the regularization term is replaced with a soft orthogonality constraint on *V*_*i*_ such that 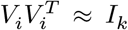 where *I*_*k*_ is the identity matrix of size *k* × *k*. We employ the NSA-Flow subalgorithm to optimize these constraints within the alternating minimization framework (Brian B. Avants, Tustison, and Stone 2025). Higher weights force the *V*_*i*_ to approach perfect orthogonality, while lower weights allow more flexibility in the learned features.

### 5.4 Impact of *D*_*k*_ (PCA vs. SVD vs. ICA)

The choice of the dimensionality reduction method *D*_*k*_ significantly influences the nature of the latent space *U*_≠*i*_. This choice affects the interpretability, computational properties, and alignment of the shared latent embedding across modalities. Below, we discuss PCA, SVD, and ICA in the context of NNHEmbed.

#### 5.4.1 PCA (Principal Component Analysis)

**Definition**: PCA computes a low-dimensional representation by projecting the data onto the top *k* eigenvectors of the data covariance matrix, maximizing the variance captured in the latent space. In NNHEmbed, applying PCA as *D*_*k*_ to the concatenated set yields the principal components corresponding to the largest eigenvalues.

##### Mathematical Formulation

For a matrix 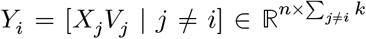, PCA solves:

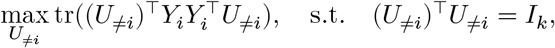

where *I*_*k*_ is the *k* × *k* identity matrix. This is equivalent to the eigendecomposition of the covariance matrix 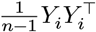.

##### Interpretation

PCA produces orthogonal components that capture the directions of maximum variance across modalities *j* ≠ *i*. In neuroimaging, for example, if *X*_*i*_ represents cortical thickness and *X*_*j*_ (for *j* ≠ *i*) represents functional connectivity, PCA ensures *U*_≠*i*_ reflects the dominant patterns of variation in *X*_*j*_*V*_*j*_, such as major connectivity networks.

##### Strengths

- Convex and computationally efficient, with a unique global solution (up to sign flips).
- Well-suited for high-dimensional data (*p*_*i*_ ≫ *n*), as it focuses on variance, reducing noise sensitivity.
- Provides interpretable components, as principal components are linear combinations of features weighted by their contribution to variance.

##### Limitations

- Assumes Gaussian-like data distributions, as it relies on second-order statistics (variance). Non-Gaussian structures (e.g., sparse neural signals) may be poorly captured.
- May prioritize variance over cross-modality alignment, potentially missing subtle shared signals.
- Sensitive to scaling differences across modalities, requiring preprocessing (e.g., centering or normalization).

#### 5.4.2 SVD (Singular Value Decomposition)

##### Definition

SVD decomposes a matrix *Y*_*i*_ = [*X*_*j*_*V*_*j*_ ∣ *j* ≠ *i*] into *Y*_*i*_ = *P* Σ*Q*^⊤^, where *P* ∈ ℝ^*n*×*k*^, Σ ∈ ℝ^*k*×*k*^ is diagonal with singular values, and 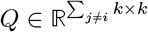. In NNHEmbed, *U*_≠*i*_ is typically set to *P*, the left singular vectors corresponding to the top *k* singular values.

##### Interpretation

SVD provides orthogonal components that maximize variance in the data, similar to PCA but applied directly to *Y*_*i*_ rather than its covariance. In NNHEmbed, SVD ensures *U*_≠*i*_ captures the principal directions of variation in the concatenated projections *X*_*j*_*V*_*j*_.

##### Strengths

- Numerically stable and unique (up to sign flips), making it robust for high-dimensional data.
- Equivalent to PCA when data is centered, providing a direct link to variance maximization.
- Computationally efficient for large matrices, leveraging optimized linear algebra libraries.

##### Limitations

- Like PCA, SVD focuses on variance, potentially missing non-Gaussian or sparse signals.
- Does not inherently account for statistical independence, limiting its ability to disentangle distinct processes (e.g., motor vs. memory signals in neuroimaging).
- Requires preprocessing to handle modality-specific scaling or noise.

#### 5.4.3 ICA (Independent Component Analysis)

##### Definition

ICA seeks statistically independent components by maximizing non-Gaussianity, often using measures like negentropy or kurtosis. In NNHEmbed, applying ICA as *D*_*k*_ to *Y*_*i*_ = [*X*_*j*_*V*_*j*_ ∣ *j* ≠ *i*] produces *U*_≠*i*_, where the columns are independent components.

##### Mathematical Formulation

ICA optimizes a contrast function, such as:

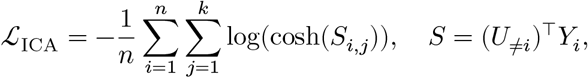

to promote non-Gaussianity in the components of *U*_≠*i*_.

##### Interpretation

ICA disentangles statistically independent signals across modalities *j* ≠ *i*. In neuroimaging, this might separate distinct neural processes (e.g., motor vs. cognitive networks) in *X*_*j*_*V*_*j*_.

##### Strengths

- Captures non-Gaussian structures, aligning with biological assumptions about sparse or independent signals (e.g., distinct brain networks).
- Robust to Gaussian noise, as it focuses on higher-order statistics.

##### Limitations

- Non-convex optimization, leading to potential local optima and sensitivity to initialization.
- Computationally intensive, as each *U*_≠*i*_ update involves a sub-optimization problem, adding complexity to NNHEmbed’s iterative process.
- Interpretability may be challenging, as independent components may not align with variance-based patterns.

#### 5.4.4 Comparative Impact on *U*_≠*i*_

- **Variance vs. Independence**: PCA and SVD prioritize variance maximization, producing orthogonal components that capture dominant patterns in *X*_*j*_*V*_*j*_. ICA, conversely, prioritizes statistical independence, which may better capture sparse or distinct signals but risks overfitting to non-Gaussian artifacts.
- **Computational Complexity**: PCA and SVD are convex and computationally efficient, with unique solutions (up to sign flips), making them suitable for large-scale data. ICA’s non-convexity introduces additional computational overhead and variability in solutions due to local optima.
- **Interpretability**: PCA and SVD yield components that are easily interpretable as directions of maximum variance, often corresponding to broad patterns (e.g., major brain networks). ICA components are independent but may be less intuitive, as they emphasize non-Gaussianity over variance.
- **Cross-Modality Alignment**: PCA and SVD may prioritize within-modality variance over cross-modality alignment, potentially missing subtle shared signals. ICA’s focus on independence can enhance cross-modality alignment by isolating shared non-Gaussian signals but requires careful tuning to avoid artifacts.
- **Application Context**: In NNHEmbed, PCA and SVD are preferred when modalities (e.g., cortical thickness, functional connectivity) exhibit strong variance-driven patterns. ICA is better suited for applications requiring separation of distinct processes (e.g., identifying independent neural signals in multi-omics or neuroimaging data).

#### 5.4.5 Practical Considerations in NNHEmbed

- The choice of *D*_*k*_ depends on the data and task. For example, in a neuroimaging dataset with *X*_*i*_ as cortical thickness and *X*_*j*_ as white matter metrics, PCA or SVD might capture broad structural patterns, while ICA could isolate specific neural processes.
- Preprocessing (e.g., centering, scaling) is critical for PCA and SVD to ensure fair contribution from all modalities. ICA requires careful selection of the non-linearity (e.g., logcosh) and initialization to stabilize results.
- NNHEmbed’s iterative optimization, where *U*_≠*i*_ is updated using *D*_*k*_, amplifies the impact of the chosen method. PCA and SVD provide stable updates, while ICA’s non-convexity may lead to variability in *U*_≠*i*_, affecting convergence and interpretability.

### 5.5 NNHEmbed Tuning

#### Parameters

Key parameters include matrix pre-processing, regularization weights and matrices and the choice of similarity measurement.

#### Initialization

NNHEmbed can be initialized randomly, using a joint SVD across concatenated modalities, or from other existing low-rank methods (e.g., RGCCA). Multiple starting points are recommended.

#### Optimization Approach

NNHEmbed uses a gradient-based alternating minimization scheme. *V*_*i*_ are updated via gradient descent steps (default optimizer is a “look ahead” approach) with soft thresholding and line search. *U*_≠*i*_ is updated via a dimensionality reduction step (PCA, SVD or ICA) on the concatenation of transformed embeddings from other modalities 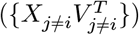.

### 5.6 Optimization and Convergence Properties

The optimization of NNHEmbed involves an iterative alternating minimization scheme, which warrants a closer look at its convergence properties and the landscape of its objective functions.

#### Objective Functions and Non-Convexity

The overall objective function is of the form:

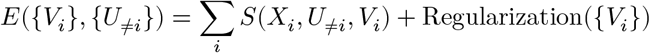

NNHEmbed employs an alternating minimization strategy over the full objective, taking advantage of analytical gradients with respect to *V*_*i*_ for each term in the function. This means it iteratively optimizes for one block of variables while holding others fixed.

- **Step 1: Optimize** *V*_*i*_ **(given fixed** *U*_≠*i*_): For a fixed *U*_≠*i*_ (and thus fixed 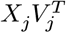 for *j* ≠ *i*), the sub-problem for optimizing a single *V*_*i*_ (e.g., for reconstruction error) becomes:

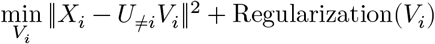

This is a standard least squares problem with regularization (e.g., L1 or L2 regularization often used with matrix factorization). When *U*_≠*i*_ is fixed, this sub-problem is **convex** with respect to *V*_*i*_. The gradient calculation presented, 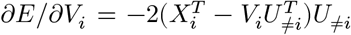, and subsequent update with thresholding (for *ℓ*_0_/*ℓ*_1_ norm regularization) and line search, aims to find the optimal *V*_*i*_ for this convex sub-problem. For the covariance forms, optimizing *V*_*i*_ given fixed *U*_≠*i*_ transforms into a series of linear updates. These steps are typically well-behaved and can be solved optimally (or sub-optimally but efficiently) for fixed *U*_≠*i*_. Analytical gradients are available for all the energies discussed, and the optimization can be performed using advanced gradient-based methodology.
- **Step 2: Optimize** *U*_≠*i*_ **(given fixed** *V*_*j*_ **for all** *j*): This step involves constructing *U*_≠*i*_ from the current 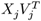 for *j* ≠ *i*. Specifically, it’s defined as *D*_*k*_(*B*) where 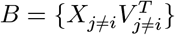 and *D*_*k*_ is a dimensionality reduction step (PCA, SVD or ICA) on the concatenation of these matrices. Given *fixed V*_*j*_ values, the transformed matrices 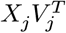 are also fixed. Concatenating them and performing SVD is a *deterministic and direct computation* (not an optimization in this step). If ICA is used for *D*_*k*_, then ICA itself involves a non-convex optimization, but *within the context of NNHEmbed’s overall loop, D*_*k*_ is treated as a “black-box” function that delivers *U*_≠*i*_ given the input *B*.

#### Convergence Properties and Guarantees

- **Local Optimum Guarantee:** The core strength of alternating minimization for non-convex problems is that if each block’s sub-problem is solved (approximately) optimally, the overall algorithm is guaranteed to converge to a *local optimum* (or at least a stationary point) of the objective function. This is because each step either decreases the objective function or leaves it unchanged. Since the objective function (a sum of squared errors or normalized covariance) is bounded below (e.g., by zero for reconstruction errors, or by −1 for normalized covariances if maximizing a similarity means bringing it close to 1), convergence is assured. The line search with *ϵ*_*i*_ explicitly ensures that only “energy improving steps” are allowed, thus guaranteeing monotonic decrease of the total energy *E*.
- **No Guarantee of Global Optimum:** Since the overall objective function is non-convex, there is no guarantee that the algorithm will converge to the global optimum. The final converged solution can be heavily dependent on the initialization strategy.
- **Sensitivity to Initialization:** NNHEmbed can be sensitive to initialization, especially with strong sparseness penalties. This is a direct consequence of the non-convex nature of the problem, leading to multiple local minima in the objective landscape. One approach is to try multiple starting points to directly addresses this limitation. However, our default strategy is to initialize the latent spaces *U*_≠*i*_ using a joint SVD across all modalities, which provides a reasonable starting point that captures the overall structure of the data.

### 5.7 NNHEmbed is Appropriate for Neuroimaging Data

NNHEmbed’s methodology provides a principled framework to directly address the complex, coupled nature of neuroimaging data:

1. **Explicit Multi-View Coupling:** Neuroimaging is fundamentally multi-view (e.g., structure, function, connectivity, genetics, behavior). NNHEmbed’s objective function inherently models how each modality’s low-dimensional representation (*X*_*i*_*V*_*i*_) can be “explained” by or is “similar” to a latent space (*U*_≠*i*_) derived from *all other modalities*. This directly reflects the notion that structural properties influence function, and functional states are enabled by structure.
2. **Reciprocal Interplay through Dynamic** *U*_≠*i*_: The “form recapitulates function” principle implies a reciprocal relationship. NNHEmbed’s dynamic construction of *U*_≠*i*_ is uniquely suited to model this. For instance, in an analysis involving structural (sMRI) and functional (fMRI) data:
  - When optimizing for *V*_*sMRI*_, *U*_≠*sMRI*_ would be derived from functional aspects (*X*_*fMRI*_*V*_*fMRI*_). This explicitly asks: “How well can my structural features be predicted/explained by the latent functional organization?” Indeed, NNHEmbed can be provided with bootstrapping or permutation approaches to estimate statistical significance for these relationships.
  - Conversely, when optimizing for *V*_*fMRI*_, *U*_≠*fMRI*_ would be derived from structural aspects (*X*_*sMRI*_*V*_*sMRI*_). This asks: “How well can my functional networks be predicted/explained by the latent structural architecture?” This inherent “mutual explanation” mechanism allows NNHEmbed to computationally explore the reciprocal dependencies that define “form recapitulates function” in a data-driven manner.
3. **Feature-Level Interpretability within a Multi-Modal Context:** The modality-specific *V*_*i*_ matrices are crucial for neuroscientific discovery. They allow researchers to identify precisely which regions, cortical thickness measures, white matter tracts, or functional networks within a given modality are most relevant to the common latent representations influenced by other modalities. This is far more interpretable than a single, merged loading matrix from concatenated approaches. For example, *V*_*sMRI*_ might reveal specific anatomical regions whose morphology is highly predictive of latent functional patterns captured by *U*_≠*sMRI*_.
4. **Flexibility in Defining Multi-Modal Relationships:** Different types of coupling may exist between neuroimaging modalities.
  - For instance, one might hypothesize that structural changes directly *reconstruct* functional activity (e.g., using a reconstruction error objective).
  - Alternatively, one might propose that latent structural and functional spaces are *linearly related* or *co-vary* (e.g., using the Canonical Covariance objective). NNHEmbed’s ability to switch between these similarity definitions allows for exploring diverse hypotheses about how brain structure and function are related, moving beyond a single model of association.
5. **Robustness to Diverse Noise Profiles:** Neuroimaging data is known for its varying noise characteristics across modalities. Structural MRI is generally high signal-to-noise and stable, while fMRI is susceptible to physiological noise, motion artifacts, and scanner instabilities. NNHEmbed’s potential robustness to view-specific noise could be highly beneficial. It mitigates the risk of one particularly noisy modality contaminating the underlying inferred latent components that are crucial for understanding the integrated brain system.

In conclusion, NNHEmbed offers a computationally elegant and neurobiologically appropriate framework for dissecting the intricate relationships between different facets of brain organization and activity. By explicitly modeling how each modality is explained by the collective information from all others, NNHEmbed provides a powerful tool to investigate the “form recapitulates function” principle, paving the way for deeper insights into brain health and disease.

### 5.8 NNHEmbed Implementation Within This Study

We evaluated NNHEmbed using *m* = 3 modalities derived from sMRI, rsfMRI, and dMRI features. Key NNHEmbed parameters and variations tested included:

- **Number of Components (k):** Fixed at 31 based on the scree plot of each data matrix in the UKB training data; 31 components explained 95% variance for the smallest input data matrix which limits the learnable rank.
- **Source Separation Function (f):** Compared Independent Component Analysis (ICA, using fastICA), Principal Component Analysis (PCA), and Singular Value Decomposition (SVD) for generating the shared basis 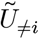 in standard NNHEmbed nomenclature.
- **Similarity Function (S):** Systematically compared:
  - **Reconstruction Loss (Reg)**
  - **Absolute Canonical Covariance Loss (ACC)**
  - **Normalized Correlation (NC)**
  - **Independence (Log-Cosh) (osh)**
- **Regularization:** Sparse regularization (*ℓ*_1_ penalty, implemented via soft-thresholding in the projected gradient descent) was applied to *V*_*i*_. Sparsity level was typically set to retain ~10-25% of features depending on the distribution and loss functions; this value was not explored and was selected based on prior work. Non-negativity constraints were enforced on *V*_*i*_ for interpretability. Alternative graph regularization was not explicitly explored in the primary analyses reported here. Additionally, we evaluated the NSA-Flow approach for imposing orthogonality constraints on *V*_*i*_ (Brian B. Avants, Tustison, and Stone 2025). The NSA-Flow method uses a flow-based approach to iteratively project *V*_*i*_ toward the Stiefel manifold, ensuring 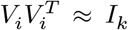 where *I*_*k*_ is the identity matrix of size *k* × *k*. This orthogonality constraint encourages distinct feature representations across the full basis and allows the sparseness to emerge naturally from the orthogonality constraint. This also eliminates the need for explicit *ℓ*_*p*_ regularization terms on the vectors within *V*_*i*_ and permits the sparseness to emerge from metrics on its global matrix structure. Of the top 20 best performing configurations, 19 used the NSA-Flow orthogonality constraint.
- **Domain Knowledge Prior:** We evaluated the inclusion of a domain knowledge prior by projecting the *V*_*i*_ onto a set of known brain networks derived from independent datasets and literature review. This amounts to a soft ridge constraint encouraging the learned components to align with known functional and structural brain networks. We compared results with and without this prior at different strengths. Of the top ten best performing configurations, six used a non-zero domain knowledge contribution. However, this component is exploratory and will require additional effort to fully evaluate its impact.
- **Parameter Combinations:** 144 combinations of mixing algorithm, similarity function, orthogonality/sparseness constraint and inclusion of prior domain knowledge were evaluated. The table of all combinations is provided in the supplementary materials.

#### 5.8.1 Evaluation Strategy

1. **Phase 1: Unsupervised Evaluation & Parameter Selection (UK Biobank):**
  - NNHEmbed models with different parameter combinations (Table 1) were trained on a large subset of UKB M3RI data (Table 4).
  - **Metrics:** Models were ranked based on feature orthogonality, RV coefficient, age and cognitive prediction and longitudinal sensitivity. The RV coefficient from permutation testing assessed the strength of the multi-view relationship captured beyond chance.
  - Based on a composite ranking incorporating these metrics, the best parameter configuration was selected for a deep dive in reporting.
2. **Phase 2: Brain-Behavior Association Validation (NNL, ADNI, PPMI):**
  - For each validation dataset, preprocessed M3RI data 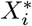 was pro-jected onto the UKB-derived feature bases: 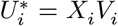. This yields *k* embedding scores per modality for each subject.
  - **Association Analysis:** Linear models were used to assess the relationship between the embedding components 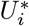 (used individually or combined) and the cognitive/functional outcome variables.
    - **Cross-sectional:** Cognitive/Functional Score ~ NNHEmbed Component(s) + Covariates (age, sex, education, brain volume/ICV). Performance measured by component p-values (fdr-corrected), effect sizes (Cohen’s *d* or *f* where appropriate), and overall model fit (*R*^2^).
    - **Longitudinal:** Using linear mixed-effects models: Score ~ Time * NNHEmbed Component(s) + Covariates + (Time | SubjectID). Assessed if component scores predicted the cognitive/functional assessments.
  - **Comparison:** Performance was compared across the different NNHEmbed configurations trained in Phase 1.

### 5.9 Cognitive and Functional Measures Used to Interpret NNHEmbed Embeddings

NNL scores were derived from the NIH Toolbox Cognition Battery (NIHTB-CB) (Weintraub et al. 2013; Carlozzi et al. 2015) and include age-corrected standard scores for individual tests as well as composite scores for cognitive domains. ADNI cognitive and functional measures included the Alzheimer’s Disease Assessment Scale-Cognitive Subscale (ADAS-Cog) (Rosen, Mohs, and Davis 1984), Clinical Dementia Rating Sum of Boxes (CDR-SB) (Morris 1993), Functional Activities Questionnaire (FAQ) (Pfeffer et al. 1982), Rey Auditory Verbal Learning Test (RAVLT) (Rey 1964), and the Mini-Mental State Examination (MMSE) (Folstein, Folstein, and McHugh 1975). PPMI measures included the Montreal Cognitive Assessment (MoCA) (Nasreddine et al. 2005) and Movement Disorder Society-Unified Parkinson’s Disease Rating Scale Part III (MDS-UPDRS III) (Goetz et al. 2008). Below are brief descriptions of key cognitive and functional measures used in this study:

#### 5.9.1 Trials (NNL)

- **Category**: Episodic Memory
- **Measures**: Immediate recall performance across learning trials.
- **Scoring**: Based on the number of items recalled per trial, with higher totals indicating stronger learning efficiency.

#### 5.9.2 Delayed Recall (NNL)

- **Category**: Episodic Memory
- **Measures**: Retention of learned material after a delay.
- **Scoring**: Number of items recalled after a set delay; higher values indicate stronger memory consolidation.

#### 5.9.3 Total Recall (NNL)

- **Category**: Episodic Memory
- **Measures**: Overall recall ability combining immediate and delayed performance.
- **Scoring**: Sum of recalled items across trials and delay periods.

#### 5.9.4 Picture Vocabulary Test Age 3+ v2.0 (NIHTB-CB; NNL)

- **Category**: Language & Crystallized Intelligence
- **Measures**: Receptive vocabulary knowledge across development.
- **Scoring**: Accuracy in identifying pictured items; higher scores reflect greater crystallized verbal ability.

#### 5.9.5 Flanker Inhibitory Control and Attention Test Age 12+ v2.1 (NIHTB-CB; NNL)

- **Category**: Executive Function / Attention
- **Measures**: Ability to suppress responses to distracting stimuli while focusing on target stimuli.
- **Scoring**: Speed and accuracy of responses; higher composite scores indicate stronger inhibitory control.

#### 5.9.6 List Sorting Working Memory Test Age 7+ v2.1 (NIHTB-CB; NNL)

- **Category**: Executive Function / Attention
- **Measures**: Capacity to store, update, and manipulate information in working memory.
- **Scoring**: Total number of correctly sorted items, with higher scores indicating better working memory capacity.

#### 5.9.7 Dimensional Change Card Sort Test Age 12+ v2.1 (NIHTB-CB; NNL)

- **Category**: Executive Function / Attention
- **Measures**: Cognitive flexibility and ability to switch between sorting rules.
- **Scoring**: Composite of accuracy and reaction time, with higher scores reflecting stronger cognitive flexibility.

#### 5.9.8 Pattern Comparison Processing Speed Test Age 7+ v2.1 (NIHTB-CB; NNL)

- **Category**: Processing Speed
- **Measures**: Speed of visual processing and discrimination.
- **Scoring**: Number of correct comparisons within a set time; higher values indicate faster processing.

#### 5.9.9 Picture Sequence Memory Test Age 8+ Form A v2.1 (NIHTB-CB; NNL)

- **Category**: Episodic Memory
- **Measures**: Ability to encode and recall temporal sequences of visual stimuli.
- **Scoring**: Accuracy in recalling the correct order of picture sequences.

#### 5.9.10 Oral Reading Recognition Test Age 3+ v2.0 (NIHTB-CB; NNL)

- **Category**: Language & Crystallized Intelligence
- **Measures**: Word reading and pronunciation ability.
- **Scoring**: Accuracy in reading printed words aloud; higher scores indicate stronger crystallized reading ability.

#### 5.9.11 Cognition Fluid Composite v1.1 (NIHTB-CB; NNL)

- **Category**: Global Cognition
- **Measures**: Composite of fluid cognitive domains (processing speed, executive function, memory).
- **Scoring**: Standardized composite score; higher values indicate better performance across fluid domains.

#### 5.9.12 Cognition Crystallized Composite v1.1 (NIHTB-CB; NNL)

- **Category**: Global Cognition
- **Measures**: Composite of crystallized abilities (language, vocabulary, reading).
- **Scoring**: Standardized composite score; higher values reflect stronger crystallized knowledge.

#### 5.9.13 Cognition Total Composite Score v1.1 (NIHTB-CB; NNL)

- **Category**: Global Cognition
- **Measures**: Overall measure of cognition combining fluid and crystallized composites.
- **Scoring**: Standardized total score; higher values indicate stronger overall cognition.

#### 5.9.14 Cognition Early Childhood Composite v1.1 (NIHTB-CB; NNL)

- **Category**: Global Cognition
- **Measures**: Broad measure of cognitive development for younger participants.
- **Scoring**: Composite of age-appropriate NIHTB measures; higher values indicate more advanced early childhood cognition.

#### 5.9.15 Rey Auditory Verbal Learning Test – Immediate Recall (RAVLT; ADNI)

- **Category**: Episodic Memory
- **Measures**: Learning ability across multiple word list trials.
- **Scoring**: Total words recalled across trials, with higher scores reflecting stronger immediate recall.

#### 5.9.16 Rey Auditory Verbal Learning Test – Forgetting (RAVLT; ADNI)

- **Category**: Episodic Memory
- **Measures**: Retention and forgetting rate.
- **Scoring**: Calculated as the difference between immediate and delayed recall, with higher values indicating greater forgetting.

#### 5.9.17 Alzheimer’s Disease Assessment Scale – Cognitive Subscale, Word Recall (ADAS-Cog Q4; ADNI)

- **Category**: Episodic Memory
- **Measures**: Word recall performance as part of ADAS-Cog.
- **Scoring**: Fewer words recalled corresponds to greater impairment.

#### 5.9.18 Modified Preclinical Alzheimer’s Cognitive Composite – Trail Making Test Part B (mPACC Trails B; ADNI)

- **Category**: Cognitive Flexibility
- **Measures**: Executive function and task switching.
- **Scoring**: Longer completion times reflect greater impairment.

#### 5.9.19 Modified Preclinical Alzheimer’s Cognitive Composite – Digit Symbol Substitution (mPACC Digit; ADNI)

- **Category**: Working Memory / Processing Speed
- **Measures**: Processing speed and executive function.
- **Scoring**: Number of correct symbol-digit pairings; lower scores indicate impairment.

#### 5.9.20 Mini-Mental State Examination (MMSE; ADNI)

- **Category**: Cognitive Aging
- **Measures**: Global cognition including orientation, attention, memory, language, and visuospatial ability.
- **Scoring**: 30-point scale; lower scores indicate impairment.

#### 5.9.21 Montreal Cognitive Assessment (MoCA; ADNI, PPMI)

- **Category**: Cognitive Aging
- **Measures**: Global cognition, sensitive to mild cognitive impairment.
- **Scoring**: 30-point scale; higher scores indicate better cognition.

#### 5.9.22 Alzheimer’s Disease Assessment Scale – Cognitive Subscale, 13-item (ADAS-Cog 13; ADNI)

- **Category**: Cognitive Aging
- **Measures**: Multiple domains including memory, language, and praxis.
- **Scoring**: Higher scores indicate greater impairment.

#### 5.9.23 Clinical Dementia Rating – Sum of Boxes (CDR-SB; ADNI)

- **Category**: Cognitive Aging
- **Measures**: Severity of impairment across six functional and cognitive domains.
- **Scoring**: 0–18 scale; higher scores reflect greater impairment.

#### 5.9.24 Functional Activities Questionnaire (FAQ; ADNI)

- **Category**: Functional Daily Living Activities
- **Measures**: Independence in instrumental daily living activities.
- **Scoring**: Higher scores indicate greater functional difficulty.

#### 5.9.25 Everyday Cognition Scale (ECog; ADNI)

- **Category**: Functional Daily Living Activities
- **Measures**: Perceived everyday cognitive functioning (patient and study partner report).
- **Scoring**: Higher scores indicate greater functional difficulty.

#### 5.9.26 Unified Parkinson’s Disease Rating Scale – Part II: Motor Experiences of Daily Living (UPDRS II; PPMI)

- **Category**: Multimodal Sensorimotor Function
- **Measures**: Motor aspects of daily living in Parkinson’s disease.
- **Scoring**: Higher scores indicate greater impairment.

#### 5.9.27 Unified Parkinson’s Disease Rating Scale – Part III: Motor Examination (UPDRS III; PPMI)

- **Category**: Multimodal Sensorimotor Function
- **Measures**: Motor symptoms and severity of Parkinson’s disease.
- **Scoring**: Higher scores indicate greater impairment.

#### 5.9.28 Postural Instability and Gait Difficulty (PIGD; PPMI)

- **Category**: Multimodal Sensorimotor Function
- **Measures**: Balance and gait-related function in Parkinson’s disease.
- **Scoring**: Higher scores indicate greater postural and gait impairment.

### 5.10 Computational Environment

The majority of analyses were performed with an Apple M2 Ultra with 24 CPU cores and 192 GB RAM using Darwin Kernel Version 24.3.0. Below we summarize the R environment and Python environment used in these analyses.

#### 5.10.1 Python Environment & Key Packages for IDP generation

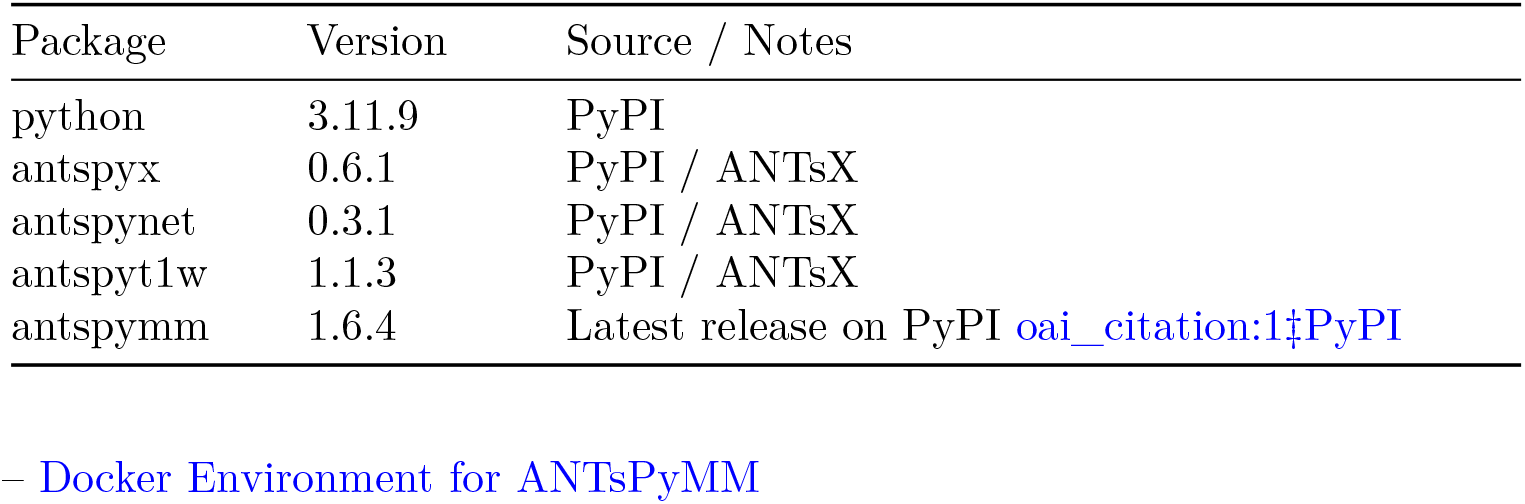

#### 5.10.2 R environment for statistical analysis

`## ### R Environment`

| Component | Value |
| --- | --- |
| R version | R version 4.4.2 (2024-10-31) |
| Platform | aarch64-apple-darwin20 |
| OS | darwin20 |

`##`

`## #### R Packages (specific to this research)`

|  | Package | Version |
| --- | --- | --- |
| ITKR | ITKR | 0.6.0.0.2 |
| ANTsR | ANTsR | 0.6.6 |
| ANTsRCore | ANTsRCore | 0.8.1 |
| subtyper | subtyper | 1.3.0 |

### 5.11 Data Availability

The data used in this study were obtained from multiple sources, each governed by specific data access and use agreements.

- **UK Biobank (UKB)**: Access to UK Biobank data requires institutional approval and acceptance of the UK Biobank data use agreement (http://www.ukbiobank.ac.uk/).
- **Alzheimer’s Disease Neuroimaging Initiative (ADNI)**: Data are available to qualified investigators through application and acceptance of the ADNI data use agreement (http://adni.loni.usc.edu/).
- **Parkinson’s Progression Markers Initiative (PPMI)**: Data access requires registration and approval via the PPMI Data and Publications Committee, with acceptance of the PPMI data use agreement (https://www.ppmi-info.org/).
- **Normative Neuroimaging Library (NNL)**: Data are available to qualified researchers upon reasonable request to the investigators, contingent upon demonstration of an appropriate research purpose and acceptance of the NNL data use agreement.

In accordance with these requirements, raw data cannot be publicly shared. However, **derived NNHEmbed solutions from this work are available at NNH feature vectors** to facilitate replication and extension of our findings.

Researchers interested in accessing the primary datasets must apply directly through the respective data access portals.

## 6 Appendix 1. Dimensionality Bounds in Joint (Multimodal) Representation Learning

In multimodal (multi-view) dimensionality reduction, each modality 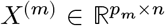 is assumed to arise from a shared low-dimensional latent representation *Z* ∈ ℝ^*k*×*n*^ through a modality-specific loading matrix 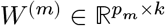:

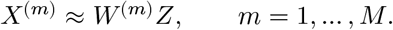

A central property of this model is that the dimensionality *k* of the shared latent space is *upper-bounded by the rank of each modality*:

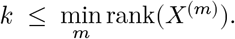

This follows from the standard linear algebra identity

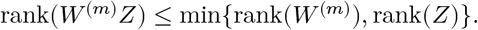

Since *W* ^(*m*)^*Z* must approximate *X*^(*m*)^, the shared latent representation cannot have rank larger than that of the data matrix it is reconstructing. Therefore, the least informative (lowest-rank) modality becomes the **bottleneck** for the joint factorization.

### 6.1 Practical Dimensionality Selection

In empirical data analysis, the *effective* rank of each modality is commonly estimated using a scree-plot criterion (e.g., the number of principal components accounting for 95% of variance). If modality *m* requires *d*_*m*_ components to reach this threshold, and

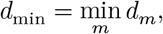

then any joint latent representation must satisfy

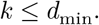

Thus, the smallest effective dimensionality among all modalities acts as an upper bound on the dimensionality of the shared embedding.

### 6.2 Extreme Case for Intuition

Consider two modalities:

- Modality A: effectively 2-dimensional,
- Modality B: effectively 31-dimensional.

Even though Modality B contains more structure, the joint factorization *W* ^(*m*)^*Z* must also represent Modality A. Because A can only support a 2-dimensional embedding, the joint latent space cannot exceed two dimensions:

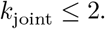

This “squeezing” effect shows that the narrowest modality controls the maximum dimensionality of the shared representation.

### 6.3 Funding

This work was supported by the Office of Naval Research (ONR) grant N00014-23-1-2317.

## Data Availability

All data produced in the present study are available upon reasonable request to the author

https://www.cohenveteransbioscience.org

https://biobank.ndph.ox.ac.uk

https://www.ppmi-info.org/access-data-specimens/data

https://adni.loni.usc.edu

## Table G: Terminology glossary

### Key terms and their definitions

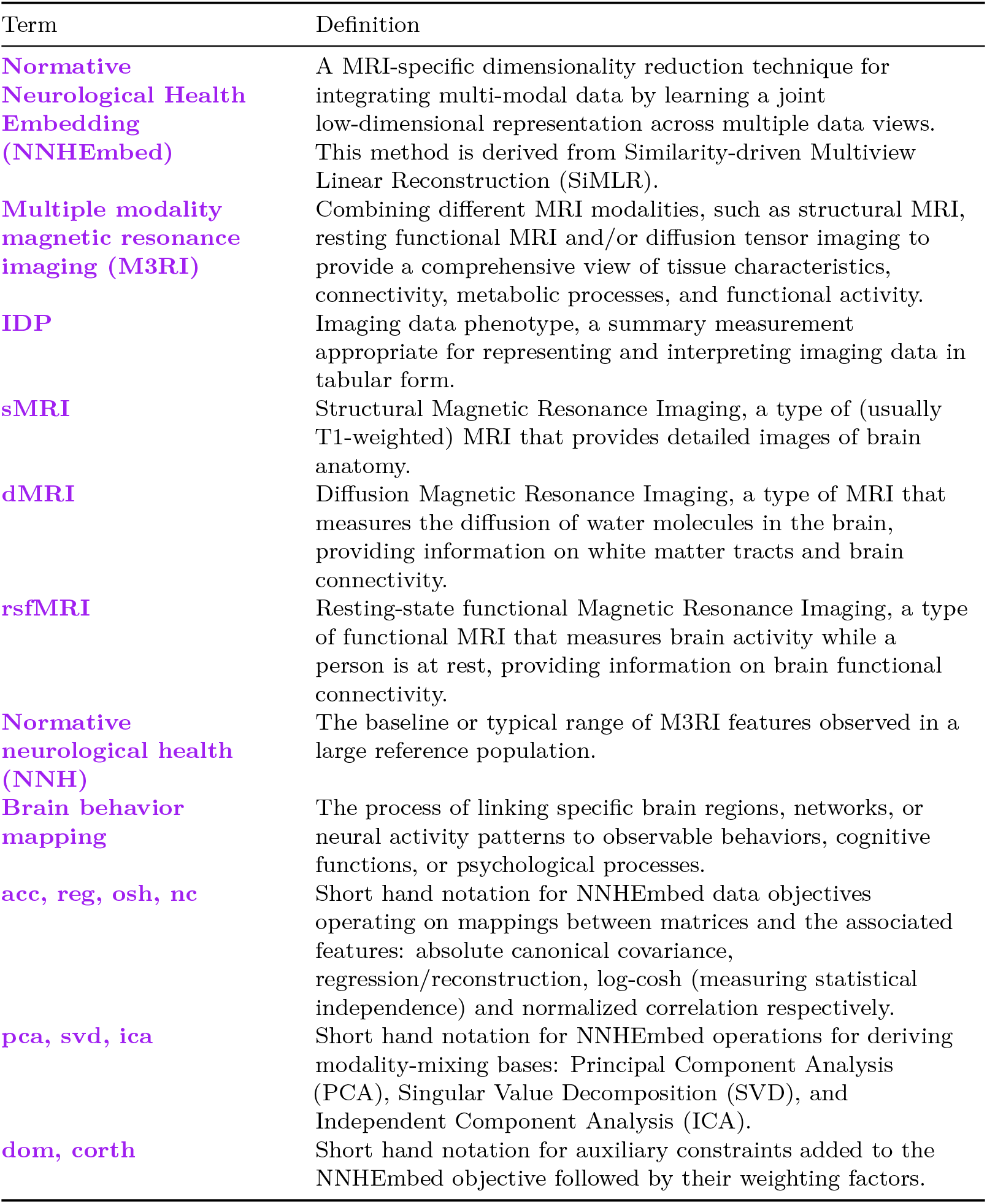

